# Bias reduction and inference for electronic health record data under selection and phenotype misclassification: three case studies

**DOI:** 10.1101/2020.12.21.20248644

**Authors:** Lauren J. Beesley, Bhramar Mukherjee

## Abstract

Electronic Health Records (EHR) are not designed for population-based research, but they provide access to longitudinal health information for many individuals. Many statistical methods have been proposed to account for selection bias, missing data, phenotyping errors, or other problems that arise in EHR data analysis. However, addressing multiple sources of bias simultaneously is challenging. Recently, we developed a methodological framework (R package, *SAMBA*) for jointly handling both selection bias and phenotype misclassification in the EHR setting that leverages external data sources. These methods assume factors related to selection and misclassification are fully observed, but these factors may be poorly understood and partially observed in practice. As a follow-up to the methodological work, we explore how these methods perform for three real-world case studies. In all three examples, we use individual patient-level data collected through the University of Michigan Health System and various external population-based data sources. In case study (a), we explore the impact of these methods on estimated associations between gender and cancer diagnosis. In case study (b), we compare corrected associations between previously identified genetic loci and age-related macular degeneration with gold standard external estimates. In case study (c), we evaluate these methods for modeling the association of COVID-19 outcomes and potential risk factors. These case studies illustrate how to utilize diverse auxiliary information to achieve less biased inference in EHR-based research.

## 1 Introduction

Electronic health record (EHR) databases allow researchers to study a wide array of diseases across patients’ entire course of medical care. However, use of observational databases such as EHR for health research presents many practical challenges that can negatively impact internal validity and external generalizability of resulting inference. Some issues include poorly measured variables, missing data, confounding, and limited information about patient recruitment mechanisms. Analytical and design-based strategies for addressing these data limitations are key to obtaining high-quality inference based on EHR data.

Researchers often are interested in using EHR data to relate a binary disease phenotype *D* to a set of predictors, *Z*, and to generalize results to a defined external target population. Two common sources of bias in these analyses are (1) lack of representativeness (selection bias) and (2) misclassification of EHR-derived disease phenotypes (information bias). Many researchers have addressed these two issues individually in the EHR setting (e.g. Haneuse and Daniels, 2016; Huang et al., 2018; Sinnott et al., 2014). However, these works do not address how to account for both sources of bias in a single data analysis. Recently, Beesley and Mukherjee (2020) proposed novel strategies for addressing these two sources of bias simultaneously. The work by Beesley and Mukherjee has several new features. Misclassification probabilities are allowed to vary across patients, and population summary statistics and internal data (e.g., patient visit patterns) are combined to address misclassification. External summary statistics or data are then used to address selection bias through weighting.

Beesley and Mukherjee (2020) demonstrates good bias reduction and inferential performance of these methods when variables related to selection (collectively, denoted *W*) and phenotype misclassification (denoted *X*) are known and observed. In reality, drivers of selection and misclassification may not be known, and known drivers may not always be observed (e.g., income, residential information, access to health care). Additionally, these methods rely on access to high-equality external data (or summary statistics) on *D* and *W* from the target population (or a probability sample). The availability of such external data will depend on the target population. External individual-level data may present additional challenges such as missing data, or we may have access to marginal distributions of variables for the target population but not their joint distribution. Our ability to correct bias will naturally be limited by the data and external information we have available. Implementation of these methods in messy real-life data analysis is not trivial, and good performance is not guaranteed by proven theoretical results. It is of interest, therefore, to explore how these methods perform for some real-world inferential problems and to provide a general road map for researchers interested in applying these methods in their own data analyses.

In this paper, we explore how the methods proposed in Beesley and Mukherjee (2020) can be applied in practical EHR data analysis through three case studies. In doing so, we demonstrate the potential for bias reduction in practice and highlight some limitations. We first consider data from the Michigan Genomics Initiative (MGI), a longitudinal EHR and genotype-linked biorepository within The University of Michigan health system. In case study (a), we examine the relationship between cancer diagnosis and gender, accounting for the *strong* enrichment of cancer patients due to ascertainment mechanisms in MGI. This case study addresses bias by leveraging cancer prevalences by age from SEER (Surveillance, Epidemiology, and End Results program by the National Cancer Institute), age distributions from the US Census, and individual-level data from NHANES (National Health and Nutrition Examination Survey by the US Centers for Disease Control and Prevention [CDC]). In case study (b), we consider the relationship between age-related macular degeneration (AMD) and several genetic loci identified by a large population-based genome-wide association study, and summary statistics for disease prevalence by age from the US CDC are used for bias reduction. Comparative gold standard results from International AMD Genomics Consortium data are available for bench-marking different bias reduction approaches. In case study (c), we apply these methods to the setting of risk factor identification for COVID-19 outcomes using data from all primary care patients tested for SARS-CoV-2 viral infection via RT-PCR at Michigan Medicine. Unlike case studies (a) and (b), we do not expect test sensitivity to vary across patients. A random sample of untested Michigan Medicine controls are used to construct weights to account for prioritized and selective testing. We compare the performance of weighting and design-based methods for evaluating and addressing selection bias due to viral testing protocols.

In **Section 2**, we introduce the two observational databases with individual-level EHR data used for our analysis. **Section 3** provides an overview of the bias-correction strategies proposed in Beesley and Mukherjee (2020). In **Sections 4-6**, we apply these methods to obtain corrected point estimates and standard errors for the three examples. We conclude with a discussion in **Section 7**. Through these case studies, our goal is to illustrate a new and valuable set of tools in the EHR data analysis toolkit and highlight important considerations to facilitate implementation.

## 2 Introducing the case studies and data sources

### 2.1 The Michigan Genomics Initiative (MGI)

The Michigan Genomics Initiative (MGI) is an EHR-linked biorepository within Michigan Medicine containing over 75,000 patients with matched genotype and phenotype information (Fritsche et al., 2018). Time-stamped ICD (International Classification of Disease) diagnosis data are available for each patient. A rich ecosystem of additional information is available for each patient, including lifestyle and behavioral risk factors, lab and medication data, geocoded residential information, socioeconomic metrics, and other patient-level, census tract-level, and provider-level characteristics.

We want to use MGI data to study the association between disease status *D* and predictors *Z* and generalize to the target US adult population. The process by which data are accumulated and the systematic differences between MGI patients and our target population must be considered to achieve this goal. **Figure A**.**1** provides a visualization of mechanisms by which patients are included in MGI. Patients are recruited among perioperative patients seen at Michigan Medicine, with targeted recruitment primarily through the Department of Anesthesiology. This naturally results in strong enrichment for diseases associated with surgical intervention, such as cancer.

We illustrate how we can address this lack of representativeness relative to the US adult population through two case studies. Case study (a) explores the relationship between gender and cancer diagnosis. Given the strong enrichment for cancer in MGI, the method for handling selection bias for this case study may have a strong impact on resulting inference. In case study (b), we investigate the relationship between age-related macular degeneration (AMD) diagnosis and 43 genetic loci previously identified as risk factors for AMD. We expect AMD diagnosis to weakly associated with inclusion in MGI after adjusting for age and other comorbidities, and the method for handling selection bias may be less impactful. These case studies are summarized in **Table 1**.

**Table 1:** Descriptions of three case studies

|  | Case study (a):<br>Cancer and Gender | Case study (b):<br>Age-Related Macular<br>Degeneration and SNPs | Case study (c):<br>SARS-CoV-2 Infection and<br>Race |
| --- | --- | --- | --- |
| *Internal Data* | Michigan Genomics Initiative | Michigan Genomics Initiative patients aged 50+ | Michigan Medicine primary care patients with viral SARS-CoV-2 test or external diagnosis |
| *Target Population* | All adults in US | US adults aged 50+ | Michigan Medicine primary care patients |
| $n$ | 40101 | 27834 | 9154 |
| $D$ | presence of cancer | presence of AMD | true SARS-CoV-2 positivity |
| $D^*$ | receipt of cancer code | receipt of macular degeneration code | positivity of first SARS-CoV-2 test or external diagnosis |
| $Z$ | gender | genotype (0/1/2) for 43 SNPs, age, genotype PCs 1-4, gender, batch | race, age, comorbidities, disadvantage index |
| Cases, (% of $n$) | 21345 (53.2%) | 1666 (6.0%) | 710 (7.8%) |
| *Sensitivity Model ($X$)* | age, number of visits, length of follow-up | age, number of visits, length of follow-up | none (assume constant sensitivity) |
| *Selection Model ($W$ and $D$)* | age, cancer diagnosis, diabetes diagnosis, coronary artery disease diagnosis, smoking, body mass index | age, AMD diagnosis, cancer diagnosis, diabetes diagnosis, coronary artery disease diagnosis | comorbidity index, age, gender, race, smoking, disadvantage index, body mass index, population density |
| *External Data* | US 2000 Census, SEER 1975-2018, NHANES 2017-18<sup>2</sup> | US 2000 Census, NIH National Eye Institute 2010<sup>2</sup> | simple random sample of untested Michigan Medicine patients<sup>2</sup> |
| *Gold Standard $\theta_Z$* | US SEER<sup>2</sup> prevalences | IAMDGC GWAS<sup>3</sup> | none |
| Assumptions<sup>1</sup> | $X^\dagger$ related to $D$ given $Z$<br>$X^\dagger$ may be indep. of $Z$ given $D$ | $X^\dagger$ may be indep. of $D$ given $Z$<br>$X^\dagger$ may be indep. of $Z$ given $D$ | sensitivity constant in $Z$ |

<sup>1</sup> $X^\dagger$ refers to the factors in $X$ (sensitivity model) not included in $Z$ (disease model). In case study (a), $X^\dagger = X$ . In case study (b), $X^\dagger$ = number of visits and length of follow-up. In case study (c), sensitivity is assumed to be constant and $X$ is empty.
<sup>2</sup> Details can be found in **Table A.2**.
<sup>3</sup> International Age-Related Macular Degeneration Genomics Consortium

In these two case studies, we consider a subset of 40,101 unrelated MGI participants (enrolled 2012-2019) who are of recent European ancestry. We first characterize some differences between this MGI dataset and our target population, the US adult population. For case study (b), we restrict our target population to US adults aged 50+. Using these data, we define observed disease variables for several phenotypes of interest (cancer, macular degeneration, coronary artery disease, and diabetes) based on whether or not patients received particular diagnosis codes during follow-up in the Michigan Medicine EHR. **Table A**.**1** provides descriptives for the patients used in our analysis and compares these in parallel to available summary statistics related to demographics and disease rates from the US adult population. **Table A**.**2** details the sources used to obtain these population summary statistics. We also provide descriptives for adults interviewed and examined for NHANES in 2017-2018, which represents a probability sample from the US adult population. We generally find that MGI patients tend to be older and have a greater burden of disease compared to patients in NHANES and the US adult population. This is expected in a hospital-based perioperative cohort. In modeling disease risk, therefore, we need to carefully address potential relationships between patient characteristics (*W*) and inclusion in MGI if we want to generalize results to the US adult population.

We will address selection bias by leveraging external summary statistics and also some individual-level data from NHANES. Since not all people in the US adult population are eligible for inclusion in MGI, we need to make implicit assumptions about transportability in adjusted disease associations (*D*|*Z*) between the MGI source and US adult populations (see **Figure A**.**1** for a visualization). However, we do not require distributions of other patient characteristics (*Z*, variables related to selection, or variables related to misclassification) to be the same between these populations, so key factors such as age and racial composition can vary between the source and target populations.

We are also concerned about the potential for bias due to misclassification in our EHR-derived phenotypes. Of the MGI patients considered, nearly 10% of patients were seen for less than 6 months and nearly 9% were seen for fewer than 10 visits. We may have little confidence in saying a person does not have a given disease if they were seen for very few visits or a very short window of time. Instead, we may have just missed the disease. Therefore, misclassification of our derived phenotypes is a strong concern, particularly given the short follow-up and small number of visits for some MGI patients.

### 2.2 COVID-19 Cohort in University of Michigan Health System

In case study (c), we consider data for 16,228 people tested or externally diagnosed with SARS-CoV-2 viral infection and seen at Michigan Medicine between March 10th and July 28th, 2020. We also have a data for a simple random sample of 30,000 untested Michigan Medicine patients. Data on race, age, body mass index (BMI), smoking status, etc. are available. Additionally, geocoded residential information was combined with US Census data to construct neighborhood-level socioeconomic factor variables. Detailed definitions for these various patient-level and neighborhood-level factors are provided elsewhere (Gu et al., 2020). We focus our attention to the 9154 tested/diagnosed patients and 4618 untested patients who received prior primary care at Michigan Medicine. We choose to focus on primary care patients due to the comparatively large amount of risk factor missingness for non-primary care patients.

Our interest is in modeling the relationship between *true* SARS-CoV-2 infection status (*D*) and patient characteristics (*Z*) among primary care patients in Michigan Medicine. It is well-known that currently used SARS-CoV-2 viral tests are imperfect, with up to 30% of infected patients receiving a false negative result (Woloshin et al., 2020). Additionally, tested patients are expected to be strongly enriched for certain respiratory and flu-like symptoms, and who gets tested may depend on other patient characteristics such as occupation (essential workers tested more often) given limited test accessibility (Allen et al., 2020). **Table B**.**1** provides a comparison between tested and untested Michigan Medicine primary care patients. Tested patients tend to be older with more comorbidities. Limitations in available data include lack of reliable COVID-related symptom data for untested patients and unavailability of occupational data for all patients. Given the data available, we want to explore how the handling of misclassification and testing impacts estimated associations between patient characteristics and infection status.

## 3 Brief overview of methods

For the sake of completeness, we briefly summarize some of the key ingredient models and methods presented in Beesley and Mukherjee (2020). We refer the reader to Beesley and Mukherjee (2020) for additional details. Let binary *D* represent a patient’s true disease status and suppose we are interested in the relationship between *D* and person-level information, *Z*. We call this the *disease model*. Let *D*^∗^ denote the EHR-derived disease phenotype, which we will assume is binary. *D*^∗^ is a potentially misclassified version of *D* with corresponding sensitivity and specificity. In this paper, we will assume specificity = 1, so *D*^∗^ is misclassified only through missed diseases. We call the mechanism generating *D*^∗^ given *D* = 1 the sensitivity model and let *X* denote patient and provider-level predictors related to sensitivity. For example, *X* may contain factors such as patient age, length of follow-up, and number of hospital visits. We suppose we model both *D*|*Z* and *D*^∗^|*X, D* = 1 using logistic regressions as follows:

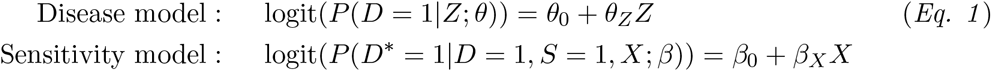

where *S* is an indicator denoting inclusion in the EHR database.

Our interest is in using the EHR data analysis to make inference about some *defined target population*. For example, we might define our target population as the US adult population between ages 50-65. This population may differ from our *source population* (e.g. people in the catchment area of the health system) (see **Figure A**.**1** for an illustration). We will assume that inference about *D*|*Z* is transportable between the source and target populations (Dahabreh and Hernán, 2019). In order to make a link between our EHR sample and our target population of interest but without loss of generality, we view our target population as if it were the source population and define a corresponding selection model describing inclusion in our EHR (*S* = 1) as a function of *D* and additional covariates (*W*), which is defined relative to the target population.

Under these modeling assumptions, Beesley and Mukherjee (2020) observes that

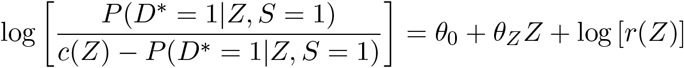

where *c*(*Z*) and *r*(*Z*) are defined as follows;

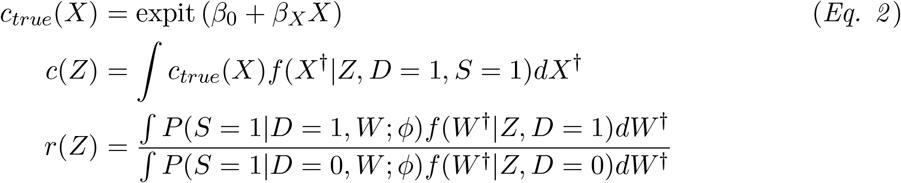

and where *X*^†^ and *W* ^†^ represent the elements in *X* and *W* not included in *Z*, respectively. The term *c*(*Z*) represents sensitivity as a function of *Z*, and *r*(*Z*) is sampling ratio as a function of *Z* and relates to the enrichment of disease in the EHR data as a function of *Z*. These two terms will rarely be known in practice, and Beesley and Mukherjee (2020) proposes a multistep strategy for estimating disease model parameters accounting for unknown *c*(*Z*) and *r*(*Z*) as illustrated in **Figure 1**. We summarize these steps as follows:

**Figure 1:**
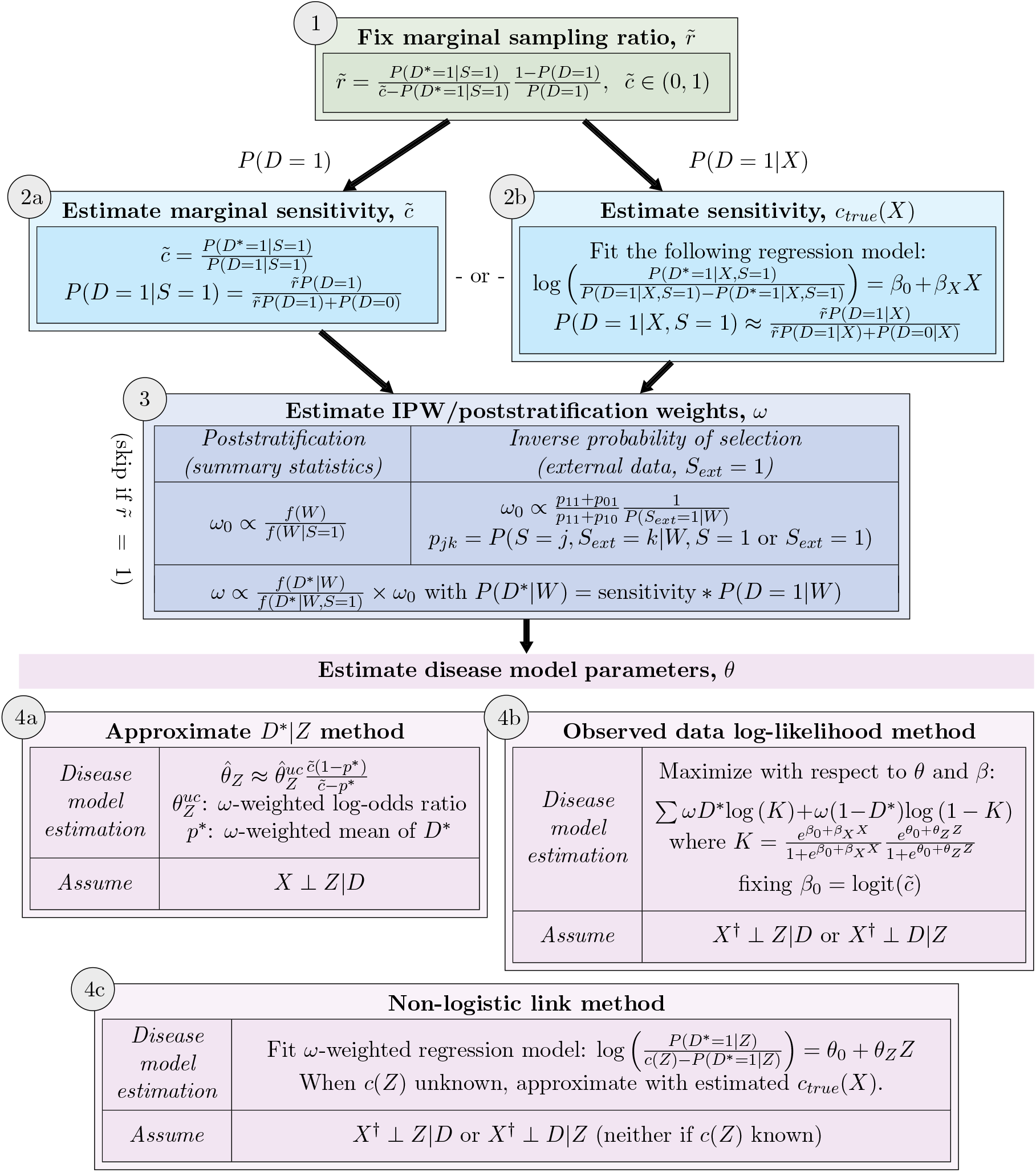
Flowchart of sensitivity and disease model parameter estimation methods ^1 2^ ^1^ <u>Notation:</u> *X*^†^: predictors in *X* (sensitivity model) not included in *Z* (disease model) *c*(*Z*): sensitivity *c*_*true*_(*X*) integrated over the distribution of *X*^†^ given *D* = 1 and *Z*. *S*_*ext*_: indicator of inclusion in external probability sample for selection bias adjustment, if available ^2^ For Step ➂ inverse probability of selection w2eighting, we set *p*_11_ = 0 if there are no subjects included in both the internal and external datasets. *p*_*jk*_ can be estimated using logistic (no overlap) or multinomial logistic (overlap) regression in the merged internal and external data. When only sampling weights are available for the external dataset, *P* (*S*_*ext*_ = 1|*W*) can be estimated using beta regression as proposed in Elliot (2009).

- <u>Step ➀: Fix marginal sampling ratio</u>, 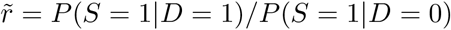. We can obtain a rough sense of 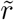 using the equation in **Figure 1**.
- <u>Step ➁: Estimate sensitivity</u>. If *c*(*Z*) is a constant, we can estimate sensitivity 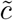 using method 2a in **Figure 1**. Otherwise, we directly estimate parameter *β* in the sensitivity model to obtain *c*_*true*_(*X*) = expit(*β*_0_ + *β*_*X*_*X*), the sensitivity as a function of *X* (via method 2b). We then approximate *c*(*Z*) with estimated *c*_*true*_(*X*).
- <u>Step ➂: Estimate weights</u>*<u>ω</u>* <u>for selection bias adjustment</u>. We consider two strategies based on the kind of information that is available in the target population. When we have individual-level data for *D* and *W* in a probability sample from the target population, we can estimate inverse probability of selection weights. When we have summary statistics of *D* and *W* available for the target population (and ideally, their joint distribution), we can obtain poststratification weights. In practice, *W* may not be available, and we will use what elements of *W* are available in an effort to *reduce* selection bias. **Figure 1** provides strategies for approximating *ω* correcting for phenotype misclassification (substituting estimated sensitivity *c*_*true*_(*X*) or 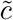 in the formula for *ω*). For comparison later on, we will also obtain uncorrected weights that ignore phenotype misclassification. These weights are calculated by setting sensitivity to 1 in the formula for *ω*.
- <u>Step ➃: Estimate disease model parameter *θ*_*Z*_</u>. **Figure 1** describes three strategies for estimating *θ*_*Z*_ under different assumptions. When sensitivity *c*(*Z*) is constant in *Z*, we can apply a simple method involving approximating the distribution of *D*^∗^|*Z* (method 4a). Two other strategies include maximization of the weighted observed data log-likelihood (method 4b) and estimation using a non-logistic link function given estimated sensitivity (method 4c).

## 4 Case study (a): association between cancer and gender using MGI

Suppose we are interested in the relationship between cancer (*D*) and gender (*Z*) in the US adult target population. MGI is strongly enriched for cancer diagnosis (53% in MGI vs a lifetime risk of 39.5% for US adults [SEER 2017]), and factors such as age, BMI, and smoking status (some of which are related to gender) are expected to be related both to cancer diagnosis and selection into MGI through other comorbidities. Therefore, there is a *strong* potential for bias due to patient selection, and the bias-adjusted results may be highly sensitive to the adjustment method. We expect the impact of misclassification to be comparatively small, since cancer history may be more routinely recorded/reported than other phenotypes. The association between cancer and gender is a convenient estimand since the direction of the association is well-understood. SEER data indicate lower lifetime cancer risk (any site, available at https://seer.cancer.gov/csr/previous.html) among women relative to men, with corresponding log-odds ratios of −0.24 (2008-2010), −0.19 (2010-2012), −0.08 (2012-2014), and −0.07 (2014-2016). This known result provides us with the opportunity to benchmark different bias reduction strategies based on the direction of the resulting cancer-gender association estimates. In addressing potential bias due to selection and misclassification in this example, we follow the four-step procedure outlined in **Figure 1** and **Section 3. Table 1** provides a detailed characterization of the various assumptions made and data used in this analysis. Here, we describe how each of these estimation steps is carried out, incorporating external summary statistics from SEER and individual-level data from NHANES.

<u>Step ➀</u>: First, we specify a value for the marginal sampling ratio, 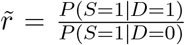, which we can view as a kind of tuning parameter roughly capturing the (unknown) degree of cancer enrichment in the study sample relative to the target population. We can use observed relationships in the data and known disease prevalence in the target population to explore plausible values of 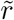 using the equation in **Figure 1** as shown in **Figure C**.**1**. To capture many potential scenarios for 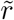 compatible with the data, we perform our analysis multiple times using the following fixed values of 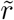: 1, 2, 5, 10, 25, 50, and 100. At the extremes, 1 corresponds to no outcome enrichment in the EHR sample, and 100 corresponds to very strong enrichment, with the probability of being included in the study sample being 100x for patients with cancer compared to patients without cancer.

<u>Step ➁</u>: Fixing 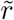, we then estimate the sensitivity with which the EHR-derived cancer phenotype *D*^∗^ captures the true cancer status *D* as a function of patient factors. We define sensitivity model covariates (*X*) to include age, the length of EHR follow-up in years, and the log-number of doctor’s visits per follow-up year. In order to perform this estimation, we use method 2b in **Figure 1**, which requires us to specify *P* (*D* = 1|*X*) from the target population. We do not know this relationship, but we do have SEER summary statistics for the relationship between age and cancer prevalence. We use these summary statistics to approximate *P* (*D* = 1|*X*) as well as we can. For some values of 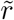 incompatible with the data, the method in **Figure 1** will provide no solution, and we focus our attention on values of 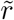 with estimable *c*_*true*_(*X*).

**Figure 2a** shows the distributions of estimated *c*_*true*_(*X*) in MGI. Median sensitivity estimates for the cancer phenotype are between 0.90 for 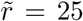 to 0.66 for 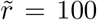. Estimated sensitivities are somewhat variable across different choices of 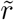 and *P* (*D* = 1|*X*) (not known), indicating a need to consider several possible values when the magnitude of the sensitivity estimates themselves are of primary interest. Previous work suggests that the downstream impact of choices for 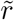 and *P* (*D* = 1|*X*) on estimated disease model parameters, however, is often small (Beesley and Mukherjee, 2020). **Figure C**.**3** provides the estimates of *β* associated with *X* in the sensitivity model. We estimate higher sensitivity with longer follow-up (years) in the EHR (log-odds ratio: 0.10, 95% CI [0.09, 0.12]) and more visits per follow-up time (log-odds ratio: 0.92, 95% CI [0.85, 1.00]).

**Figure 2:**
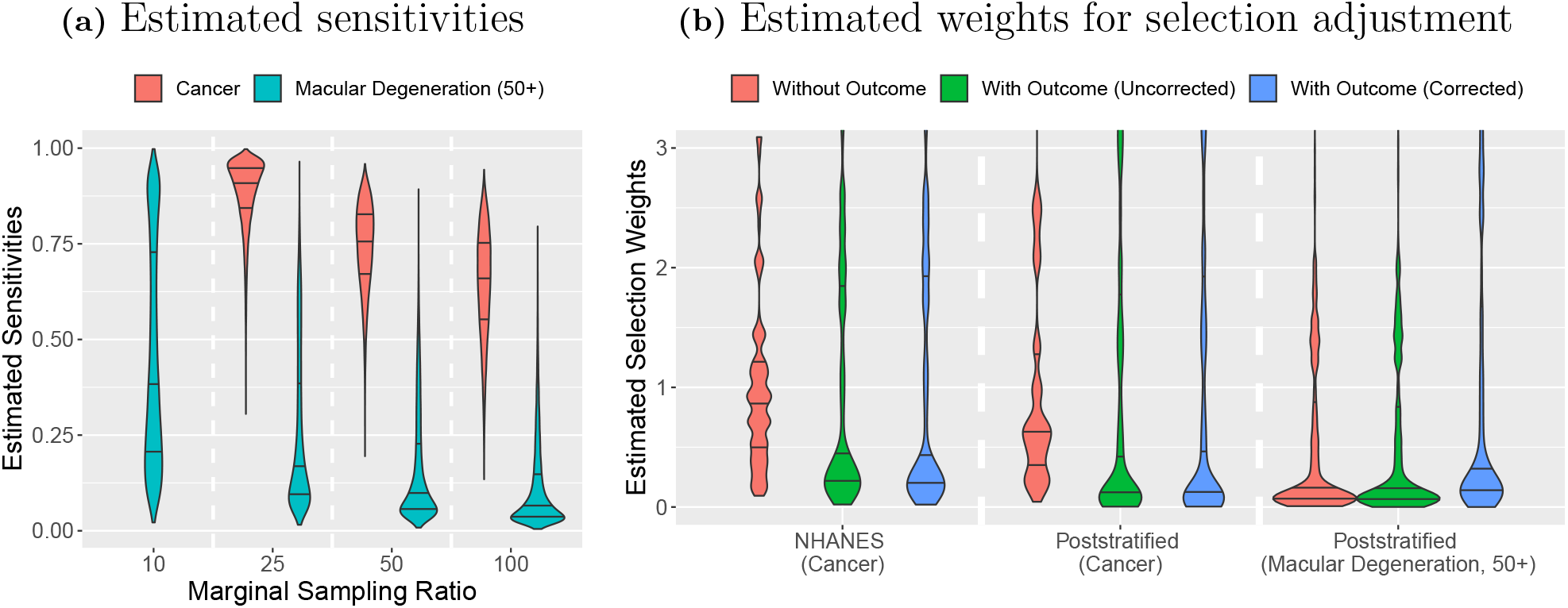
Estimated sensitivities and selection adjustment weights [Case studies (a) and (b)]^1^ ^1^Corrected weights using 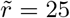 are shown. All weights were trimmed at 10. Horizontal black lines correspond to the 25th, 50th, and 75th quantiles

<u>Step ➂</u>: We consider two different strategies to account for selection bias given fixed sensitivity estimates. In the first strategy, we use summary statistics from SEER, the US Census, and the US CDC to construct poststratification weights. In the second strategy, we use publicly-available data from the NHANES (2017-2018) to construct inverse probability of selection weights. In constructing these weights, our goal is to account for some of the systematic differences between patients in MGI and patients in the US adult population. In **Table A**.**1**, we demonstrate that MGI is enriched for patients with more comorbidities (e.g. diagnosis of coronary artery disease [CAD] or diabetes), and MGI patients tend to be older than the average adult in the US. This information is incorporated into our adjustment for selection bias (included in *W*). In this section, we provide details on how these weights were estimated.

Pulling summary statistics from various sources as in **Table A**.**2**, we first define three varieties of poststratification weights. Since the joint distribution of various disease diagnoses given age is not readily available for the target population, we incorporate multiple disease diagnoses assuming independence given age. First, we define weights estimated *ignoring the cancer outcome* as follows:

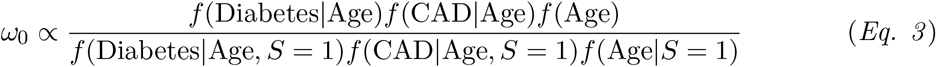

where “Diabetes”, for example, corresponds to an indicator for whether the patient received a diabetes diagnosis. The distributions in the numerator are obtained using population summary statistics, and distributions in the denominator are estimated using MGI data. We then *incorporate cancer diagnosis* into the weight estimation while correcting for phenotype misclassification as follows:

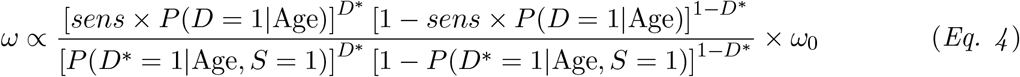

where *sens* is estimated sensitivity (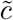 or *c*_*true*_(*X*)). We substitute population summary statistics (numerator) and MGI estimates (denominator) to obtain these weights. For comparison, we also obtain weights ignoring misclassification by setting *sens* = 1.

To compare weights estimated using different external data sources, we also obtain inverse probability of selection weights using individual-level data from NHANES, incorporating additional information about smoking status and body mass index (BMI). Let *S*_*ext*_ = 1 refer to inclusion in NHANES and *S* = 1 refer to inclusion in our MGI data. We will assume no patients are included in both databases. We first estimate weights *ignoring the cancer outcome* as follows:

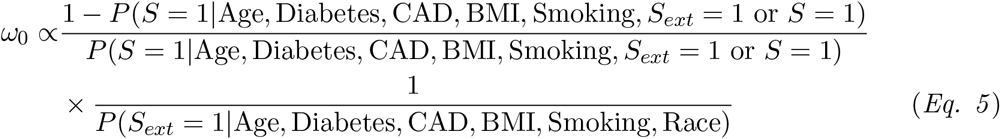

The first term accounts for differences between MGI and NHANES and is estimated using logistic regression modeling in the combined MGI and NHANES data. The second term accounts for differences between NHANES and the US adult population. Since NHANES selection weights are provided (but not the selection models themselves), we model NHANES selection using beta regression on the inverted NHANES selection weights (Elliot, 2009). These logistic and beta regression estimates are provided in **Table C**.**1**.

To obtain weights that incorporate cancer diagnosis and also account for phenotype misclassification, we multiply *ω*_0_ from *Eq. 5* by the following:

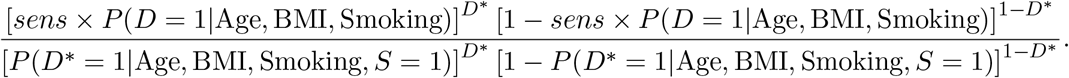

We obtain population *P* (*D* = 1|Age, BMI, Smoking) by fitting a regression model in the NHANES data weighted using the NHANES sample weights. For comparison, we again obtain weights that do not correct for misclassification by setting *sens* = 1.

Estimated poststratification weights and NHANES-based inverse probability of selection weights (after scaling to sum to the number of patients in MGI) are shown for 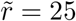 in **Figure 2b**. Other values of 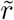 are similar. We can see substantial differences in the distribution of weights that do and do not incorporate the cancer outcome. Additionally, weights obtained using NHANES and using SEER/US Census summary statistics tend to be fairly similar in terms of their overall distributions. Weights obtained using these two methods, however, can sometimes differ substantially within individual patients.

<u>Step ➃</u>: Given estimated sensitivity and selection weights, we apply the methods in **Figure 1** to estimate the association between cancer and gender (reference = male). Results are shown in **Figure 3** and **Table C**.**2**. Uncorrected analysis results in an estimated log odds ratio of −0.07 (95% CI −0.11, −0.03). When we account for misclassification but not selection using different methods (4a-c in **Figure 1**), we see little qualitative differences in point estimates across methods. This may be due to the fairly high estimated sensitivities for the EHR-derived cancer outcome. Additionally, it may be reasonable to assume that gender (*Z*) is independent of *X* given *D*, so sensitivity *c*(*Z*) may be viewed as constant in *Z*. Assumptions for all three misclassification adjustment methods are satisfied in that case. Interestingly, estimated confidence intervals are narrower for the non-logistic link method (patient-varying sensitivity, interval width: 0.091) than for the approximation method (marginal sensitivity, interval width: 0.122) when we only account for misclassification. This small efficiency gain comes from incorporating covariates *X* related to *D* into sensitivity estimation.

**Figure 3:**
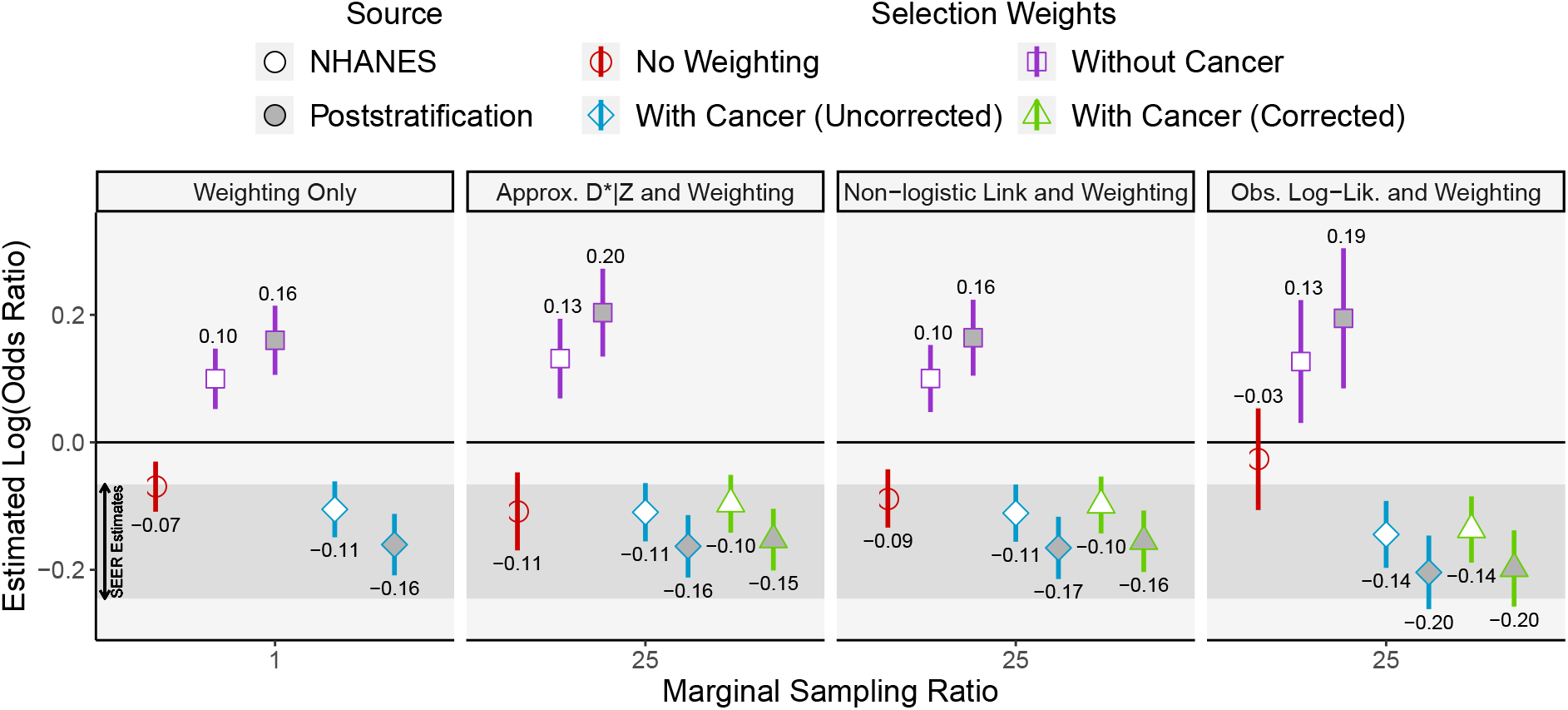
Cancer-gender log-odds ratio after applying proposed selection weighting and mis-classification adjustment methods (reference = male) [Case study (a)]^1^ ^1^ Results using a marginal sampling ratio of 25 are shown. Results for sampling ratios of 50 and 100 are similar. The horizontal shaded region corresponds to the range of SEER estimates using data between 2008 and 2016. “Approx. *D*^∗^ *Z*”, “Non-logistic Link”, and “Obs. Log-Lik.” correspond to methods 4a, 4c, and 4b in **Figure 1**, respectively. The log-odds ratio estimate is printed near each plotted confidence interval.

We see large differences in the estimated log-odds ratios when we use different selection weights to account for selection bias. In particular, weights excluding the cancer diagnosis outcome produce point estimates in entirely the “wrong” direction (e.g. a log-odds ratio of 0.09, 95% CI: [0.05,0.15]), reflecting the strong need to incorporate the direct impact of cancer diagnosis on selection when specifying the weights. When we incorporate the cancer outcome in constructing the weights, the resulting point estimates are in the “right” direction (indicating lower rates of cancer diagnosis in women compared to men) for both the NHANES and poststratification weighting strategies (e.g. −0.10, 95% CI: [-0.14,-0.05] for NHANES IPW and −0.15, 95% CI: [-0.20,-0.10] for poststratification under approximation method). Additionally, we obtain narrower confidence intervals when we account for selection bias using weights that incorporate the outcome relative to weights that do not incorporate the outcome (e.g. widths 0.108 vs 0.096 for poststratification weighting without misclassification adjustment). In this example, we see little impact of correcting for phenotype misclassification in weight development, perhaps due to the high estimated sensitivities for the cancer phenotype. The estimated log odds ratios differ somewhat for weights obtained using poststratification vs. NHANES, where poststratification produced stronger cancer-gender associations. This result serves as a cautionary tale against ignoring the outcome when estimating selection bias adjustment weights when the outcome is strongly related to selection. Additionally, we get somewhat different results when selection is addressed using different external data sources, and researchers may want to compare results using several different sources in practice.

## 5 Case study (b): association between macular degeneration and genetic loci using MGI

We now estimate associations between previously identified genetic loci and age-related macular degeneration (AMD) diagnosis using MGI data, adjusting for other patient factors such as age at last visit, gender, and principal components of the genotype data. We define our target population as the US adult population aged 50+ (**Table 1**). AMD is weakly enriched in MGI relative to adults aged 50+ in the US population (**Figure A**.**2**), and we may expect individual genetic loci in *Z* to be at most weakly associated with selection. Therefore, we may be less concerned with handling of selection bias in this example compared to case study (a). Additionally, we hypothesize that underreporting of disease may be a bigger challenge for case study (b), since we may expect many patients are treated for AMD through local health care providers, and consequently a large number of AMD diagnoses may be missed in the Michigan Medicine EHR. These missed diagnoses may strongly impact estimation of genetic associations. In this second example, we focus on 43 independent genetic loci identified with small p-values (*<*5×10^−8^) in a genome-wide association study of over 16,000 advanced AMD cases and 18,000 controls using International AMD Genomics Consortium (IAMDGC) data (Fritsche et al., 2016). Across these 43 loci, MGI and IAMDGC GWAS log-odds ratio point estimates have a Lin’s concordance correlation coefficient (CCC) of only 0.55, and uncorrected MGI point estimates generally tend to be closer to the null compared to the IAMDGC estimates (**Figure C**.**5**). The “winner’s curse” resulting in inflated IAMDGC point estimates explains some differences, but bias due to selection and misclassification in MGI may also contribute. Below, we apply our methods to explore the extent to which systematic differences in GWAS results between these two studies may be corrected by addressing phenotype misclassification and potential selection bias.

<u>Step ➀:</u> As with case study (a), we perform our analysis in Steps 2-4 across different values of the tuning parameter, 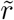, between 1 and 100.

<u>Step ➁</u>: Fixing 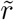, we estimate sensitivity as a function of patient-level covariates using the same method as in case study (a). Results are shown in **Figure 2a**. Sensitivity of the macular degeneration phenotype is estimated to be generally much lower than in case study (a) across all values of 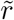, with median sensitivity ranging between 0.38 for 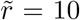 and 0.06 for 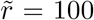. Higher sensitivities for the cancer phenotype may be related to a more complete disease history for cancer diagnoses relative to macular degeneration diagnoses as entered into the EHR through diagnosis codes.

<u>Step ➂</u>: We use summary statistics from SEER, the US Census, and the US CDC to construct poststratification weights. For this outcome, we define weights as if the target population were all US adults. Our analysis then uses data and weights from only the MGI patients aged 50+, with weights re-scaled to sum to the number of patients aged 50+ in MGI. We obtain three varieties of poststratification weights for the AMD outcome. First, we define weights *ω*_0_ *ignoring the AMD outcome* as in *Eq. 3* except this time we also incorporate the association between cancer diagnosis and age into both the numerator and denominator. We include cancer diagnosis in the weight definition to account for the strong association between cancer diagnosis and inclusion in MGI, but we do not account for misclassification of the cancer phenotype for this analysis. We then define weights that *incorporate the AMD outcome* using *Eq. 4*, where this time *D* and *D*^∗^ correspond to the AMD outcome and *sens* is either the estimated sensitivity (correcting for misclassification) or *sens* = 1 (ignoring misclassification). Resulting weights are shown in **Figure 2b** for 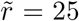. Other values of 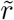 are similar. Unlike case study (a), weights that do and do not incorporate the AMD outcome tend to have similar distributions, reflecting a comparatively small impact of AMD on the probability of inclusion in MGI.

<u>Step ➃</u>: We then apply the methods in **Figure 1** to obtain bias-corrected point estimates relating macular degeneration diagnosis to 43 genetic loci in MGI. The differences between IAMDGC and MGI point estimates across loci are characterized using three metrics: (i) average absolute difference across 43 pairs of estimates, (ii) Lin’s concordance correlation, and (iii) the average absolute percent difference between the MGI and the IAMDGC estimate, relative to the IAMDGC estimate (denoted MAPE; mean absolute percentage error). We also present the average estimated MGI standard errors relative to IAMDGC. Results for the best-performing methods are summarized in **Table 4a**. Results for other methods can be found in **Table C**.**3**. We present results using 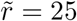, but other 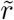 values with estimable sensitivity (10, 50, 100) are similar. When we correct for selection *or* misclassification, Lin’s concordance correlation mea-sure increases from 0.55 (uncorrected) to 0.68 (corrected). Correcting for both misclassification and selection bias did not produce additional improvement for this metric (Lin’s of 0.70). Analyses that accounted for selection (with or without misclassification bias adjustment) resulted in increased (worse) MAPE relative to uncorrected analysis (range 0.87-1.21 vs. uncorrected MAPE of 0.84). Unweighted analyses accounting for misclassification but not selection produced similar or better MAPE compared to uncorrected analysis. All bias-correction strategies shown in **Table 4a** result in point estimates that are closer to IAMDGC point estimates than in uncorrected analysis on average. Overall, the method approximating the *D*^∗^|*Z* distribution with no selection bias adjustment performs the best among the methods considered in terms of similarity between bias-corrected estimates and IAMDGC estimates. Since selection seems to be at most weakly associated with AMD diagnosis, it is not surprising that methods without selection adjustment generally perform well. Analyses incorporating selection weights had larger standard errors without much gain in terms of bias adjustment, suggesting that selection weighting did not improve inference.

**Table 4b** compares the ranked p-values for each of the 43 genetic loci after bias adjustment to the ranking in IAMDGC. Among the top 5 associations in IAMDGC, the majority are also identified as top associations in MGI. P-values produced by bias correction methods accounting only for misclassification but not selection (no weighting) tend to produce p-values very close or even identical to uncorrected analysis. In Beesley and Mukherjee (2020), we demonstrate that p-values from the non-logistic link function method (ignoring selection) will only differ substantially from uncorrected analysis when *X*^†^, representing the factors driving sensitivity not adjusted-for in the disease model, is associated with *Z* given *D*. We may be less concerned about the impact of misclassification on p-values when these terms are at most weakly associated (as in this case study). Once selection bias adjustment is incorporated, however, the resulting p-values are impacted, as seen in **Table 4b**. In general, selection may often be ignorable when estimating associations with genetic loci. However, we recommend comparing analyses with and without weighting in settings when selection may be more strongly related to *Z*.

## 6 Case study (c): accounting for selective and imperfect diagnostic testing for modeling COVID-19 susceptibility

We consider data for 9154 Michigan Medicine primary care patients tested for SARS-CoV-2 virus or externally diagnosed with COVID-19. We are interested in estimating the adjusted association between infection and race/ethnicity, accounting for potential misclassification of viral test results and the lack of representativeness of the tested patients relative to our Michigan Medicine target population. The association between race and confirmed viral infection is complicated to evaluate, since race could be related to rates of testing and/or infection rates. A careful approach is needed to tease apart associations with race using the available data on who was tested and who tested positive. For this analysis, we roughly group race/ethnicity into three categories: non-Hispanic White (NHW), non-Hispanic African American (NHAA), and Hispanic/multi-racial/other.

We first consider the question of misclassification of RT-PCR viral tests. Unlike case studies (a) and (b), misclassification corresponds to results of a diagnostic assay and is not expected to be related to patient-level factors. Recent research provides insight into plausible values for sensitivity and specificity of these test results, where specificity is generally very high (we will assume specificity = 1) and sensitivity is believed to be between roughly 0.70 and 0.90 (Woloshin et al., 2020). In the setting where *sensitivity is a constant*, we can apply the method 4a in **Figure 1** to account for imperfect test sensitivity. We note that the multiplicative constant,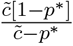 will be close to 1 unless *p*^∗^, which represents the theoretical test positive rate in the population, is large. If the population disease rate is low (say, *<*5%), the impact of imperfect test sensitivity will be small. Assuming a low population infection rate, we can *ignore the outcome misclassification* and instead focus our attention on correcting potential selection bias. Estimation addressing both sources of bias produces very similar results (not shown).

We use data from a *simple random sample* of 4618 untested Michigan Medicine primary care patients to construct weights for selection bias adjustment. Unlike the previous case studies, we have individual-level data that is representative of the target population, and we can directly model differences between the tested patients and the target population. For simplicity, we will assume that testing (so, *S*) is independent of true infection status (*D*) given available covariates (*W*), where *W* includes patient-level factors of age, number of comorbidities (e.g. liver disease, kidney disease), gender, race, smoking status, BMI and neighborhood-level factors of population density and neighborhood disadvantage index. Since testing practices changed substantially across calendar time, we model testing separately for tests from March 10th-31st (quarter Q1) and from April 1st-July 28th (quarters Q2 and Q3). Details about weight estimation including handling of missing data can be found in **Section D**. Parameters in the models for testing associated with race/ethnicity are shown in **Figure 5**. We estimate higher rates of testing among NHAA patients relative to NHW patients in quarter 1 (when testing capacity was limited), and we see no difference in adjusted testing rates between people with Hispanic/Other race/ethnicity and NHW patients. In quarters 2 and 3 (when testing capacity became less limited), the testing rates by race change, with NHW patients having similar testing rates to NHAA patients but higher testing rates than people identifying as Hispanic/other/multi-racial.

**Figure 4:**
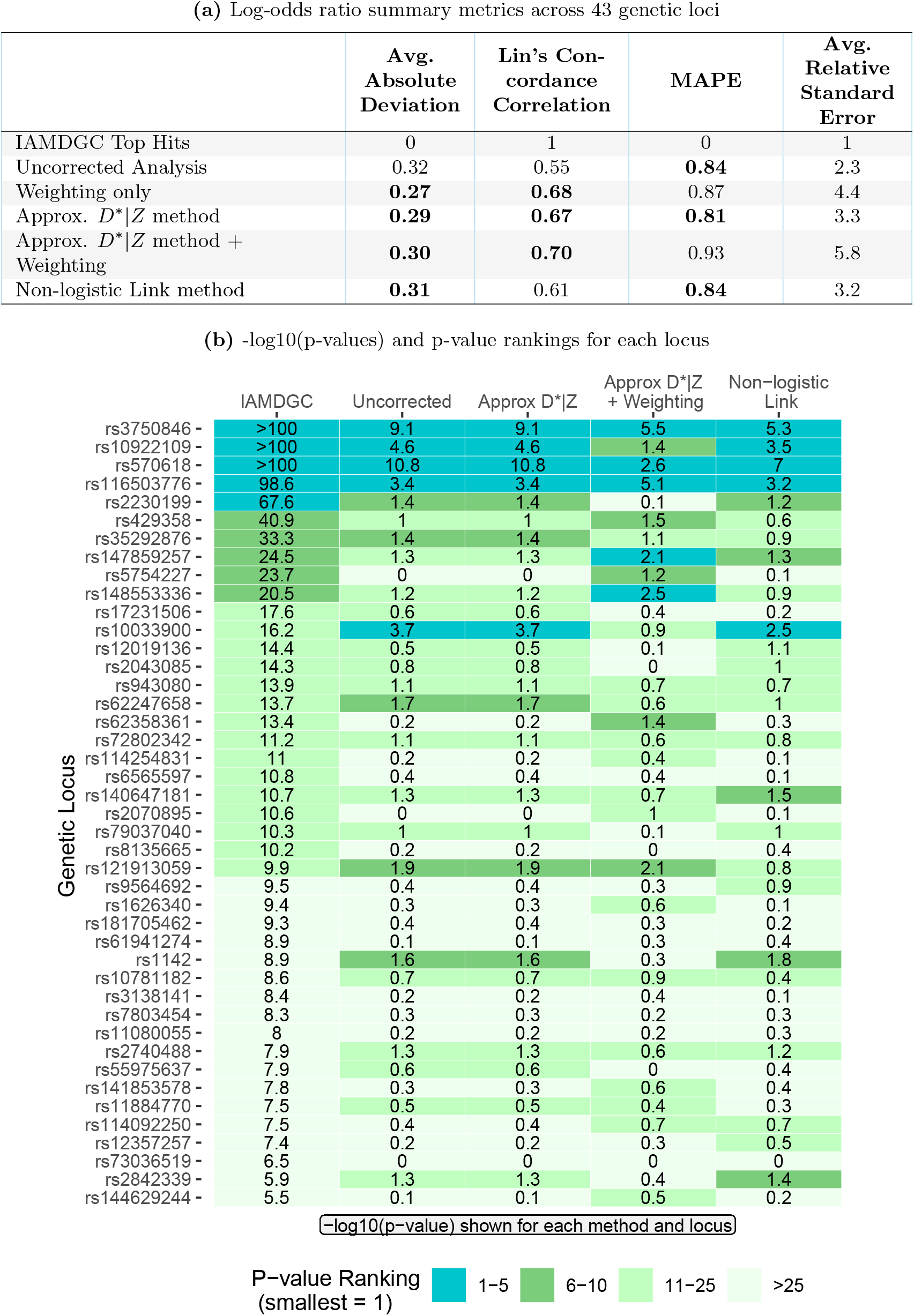
Bias-adjusted AMD log-odds ratios across 43 genetic loci and corresponding p-values [Case study (b)]^1^ ^1^For Approx. *D*^∗^ *Z* [method 4a] and Non-logistic link function [method 4c] strategies, sensitivity is estimated assuming 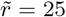. Methods with weighting used weights *ignoring the AMD outcome*. Bolded values indicate the best performing methods. **Definitions:** Average absolute deviation = average absolute difference between MGI and IAMDGC point estimates (lower is better); Lin’s concordance correlation = estimated concordance between MGI and IAMDGC point estimates (higher is better); MAPE (mean absolute percentage error) = average absolute difference between 1 and the ratio of MGI and IAMDGC point estimates (lower is better); Avg. relative standard error = ratio of standard errors for MGI and IAMDGC point estimates

**Figure 5:**
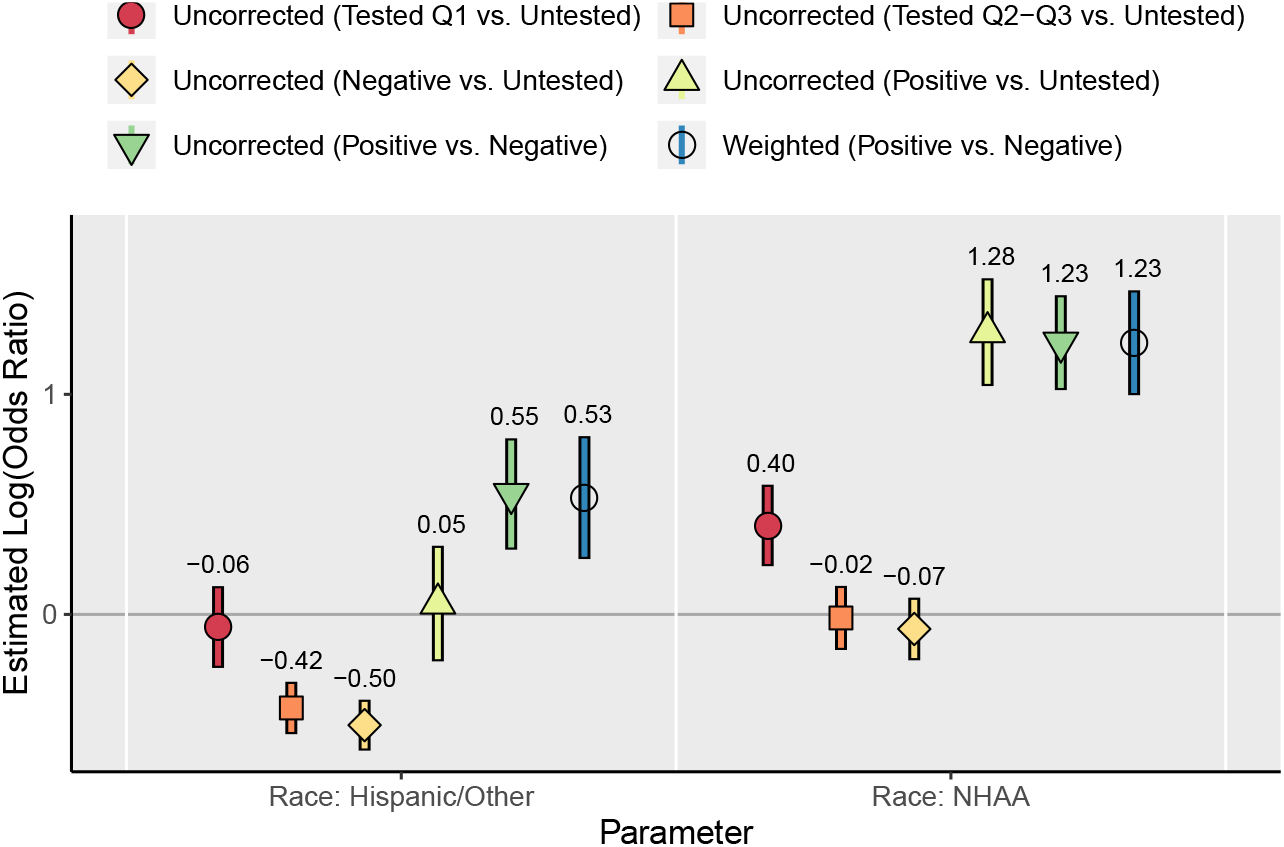
Associations between race and coronavirus testing and infection rates (reference = Non-Hispanic White) [Case study (c)]^1^ ^1^ Parentheses in the legend labels correspond to the binary outcome comparison. For example, negative vs. untested corresponds to a model for testing (yes/no) excluding test-positive patients. NHAA = Non-Hispanic African American. Q1 corresponds to testing between March 10th and March 31st, 2020. Q2-Q3 corresponds to testing between April 1st and July 28th, 2020. Weights adjusted for age, race, gender, number of comorbidities, BMI, neighborhood disadvantage index, smoking status, and population density. The log-odds ratio estimate is printed above each plotted confidence interval. Details about specification of *Z* and *X* can be found in **Table 1**.

After estimating weights to (at least partially) account for testing/selection bias, we estimate disease model parameters by fitting weighted logistic regression models for receipt of a positive vs. negative test within the tested cohort, where predictors *Z* include race, age, number of comorbidities (e.g. liver disease, kidney disease), neighborhood disadvantage index, gender, and population density. **Figure 5** shows the resulting point estimates for race. Other model parameters are shown in **Figure D**.**2**. We generally see little difference between weighted and unweighted analysis. We find that NHAA patients tend to have higher rates of viral infection compared to NHW patients (weighted log-odds ratio 1.23, 95% CI: [1.02, 1.45]) as do people with Hispanic/other race (0.55, 95% CI: [0.29, 0.80]). These estimated associations are qualitatively similar to those reported in other recent studies, including Allen et al. (2020) and Chadeau-Hyam et al. (2020).

Weights for selection bias adjustment were estimated accounting for patient factors such as age, BMI, and comorbidities, but conspicuously missing from our models for testing is information about COVID-19-related symptoms (e.g. cough, fever) or patient occupation, which were unavailable for some or all patients in our study. Since symptoms and occupation are strongly related to testing rates (particularly for tests early in 2020), we may question how well these weights can account for potential selection bias. Instead, we might appeal to design-based approaches to evaluate the potential impact of testing on disease model estimates. When we model test positivity among tested patients, we are treating patients that test negative as controls. This strategy is known as a “test-negative” design Vandenbroucke and Pearce (2019). One limitation of this approach is that patients who test negative were tested for a reason; they may have other respiratory symptoms, work an essential job requiring testing, or be at greater risk of adverse coronavirus-related outcomes, and these people will likely not represent the target population (Allen et al., 2020). In the literature for test-negative designs, researchers evaluate the potential for bias due to lack of representativeness by comparing analysis results to associations between test positive patients or test negative patients and untested population controls. **Figure 5** provides estimated associations between race and positive or negative test status using untested Michigan Medicine patients as controls. NHAA race was associated with a higher rate of positive test results (log-odds ratio 1.28, 95% CI: [1.04, 1.53]) but not negative test results (log-odds ratio −0.07, 95% CI: [-0.21, 0.08]) compared to the untested patients, supporting that NHAA patients may have higher infection rates. This analysis provides an example where design-based strategies may provide better tools for evaluating and correcting for the impact of testing relative to weighting strategies when high-quality data on important determinants of testing are unavailable.

Researchers may often be interested in modeling post-diagnosis outcomes such as hospitalization or ICU admission among patients testing positive for viral infection. We may often be less concerned about the potential impact of selection bias and misclassification when we are studying post-diagnosis outcomes, since these comparisons would often condition on having tested positive. We demonstrate this phenomenon in **Figure D**.**3**, where we apply the weights obtained previously to a model for hospitalization among people testing positive. Weighted and unweighted estimates are very similar. We find that NHAA test-positive patients tend to have higher rates of hospitalization compared to NHW test-positive patients (weighted log-odds ratio 0.86, 95% CI: [0.27, 1.46]) as do people with Hispanic/other race (0.86, 95% CI: [0.18, 1.53]). These estimated associations are consistent with those reported in Gu et al. (2020).

## 7 Discussion

Many statistical challenges arise in the analysis of electronic health record (EHR) data, including limitations in data quality (i.e., measurement error, missing data, etc.), lack of representativeness (i.e., who is in the study?), and generalizability (i.e., what do results say about my target population?). In Beesley and Mukherjee (2020), we proposed a suite of statistical tools for addressing measurement error and selection bias in disease modeling using EHR data. That work demonstrated good performance of the proposed methods when key factors related to selection and measurement error are observed, but these driving factors may be unknown or only partially measured in practice. In this paper, we explore how these statistical bias-correction and inference strategies perform in real-world data analysis through three EHR data analysis case studies. We emphasize that the goal of *unbiased* estimation in EHR data analysis is unrealistic given limitations in data availability and many competing sources of bias. Instead, our goal in implementing these methods is to produce *less biased* inference.

In the first two case studies, we consider data from the Michigan Genomics Initiative, a longitudinal EHR-linked biorepository effort within Michigan Medicine. For both of these case studies, comparative gold standard disease associations were used to benchmark the performance of various bias reduction strategies. In case study (a), bias-corrected point estimates for the association between cancer and gender were consistent with associations reported by SEER as long as the cancer outcome was incorporated into development of selection weights. In case study (b), these bias reduction methods resulted in point estimates closer on average to previously identified associations than uncorrected analysis (Fritsche et al., 2016). These case studies demonstrate that the bias correction and inference strategies from Beesley and Mukherjee (2020) may be useful for *reducing* bias in EHR-based studies even when factors related to selection and misclassification are not well-understood or fully measured. Additionally, these examples highlight the need to tailor the statistical approach to the problem at hand and illustrate settings where disease model inference can be sensitive to our strategy for handling bias adjustment.

In the third case study, we explore the very timely and important challenge of risk factor evaluation for SARS-CoV-2 viral infection. When modeling viral infections using test-negative patients as controls, we may not expect misclassification to appreciably impact absolute logistic regression parameter estimates unless the true rate of disease is substantial (e.g. *>*5%). Instead, we suggest researchers should focus on accounting for potential bias due to enhanced testing in certain patient populations, which could have a stronger impact on disease model parameters. This example highlights a setting where design-based strategies for evaluating and addressing selection biases may outperform weighting strategies when high-quality data on key factors related to selection/testing in untested patients are unavailable.

This work provides a roadmap for practical implementation of the methods for handling phenotype misclassification and selection bias in EHR data analysis proposed in Beesley and Mukherjee (2020). These methods are summarized in **Figure 1**, and Steps 2 and 4 can be easily implemented in R using package *SAMBA* available at https://cran.r-project.org/web/packages/SAMBA/index.html. These methods rely on an assumed logistic regression structure for the distribution of *D*^∗^ given *D* = 1 and covariates *X*. When potential *X* has large dimension, penalization methods could be incorporated to aid in estimation of *β*. Estimation of weights for selection bias adjustment (Step 3) presents a harder problem, and several strategies for estimating these weights are highlighted in **Figure 1**. When external individual-level data from the target population are available, inclusion in the EHR sample (i.e., *P* (*S* = 1|*W, S* = 1 or *S*_*ext*_ = 1)) can be directly modeled. In case studies (a) and (c), we use logistic regression to model this selection probability in the merged internal and external datasets, but more sophisticated modeling strategies, penalization, etc. can also be used to estimate these probabilities and construct selection weights. Additional strategies for handling multi-stage sampling, overlapping EHR and external probability samples, and use of probability samples from a different target population can be found in Beesley and Mukherjee (2020).

## Data Availability

Michigan Genomics Initiative data are available after institutional review board approval to select researchers. See https://precisionhealth.umich.edu/our-research/michigangenomics/ for details.

https://cran.r-project.org/web/packages/SAMBA/index.html

## Acknowledgments

We thank Chad Brummett, Goncalo Abecasis, and Sachin Kheterpal along with many collaborators and staff at MGI and MGI participants for donating their biosamples for research. This work was supported by The University of Michigan Comprehensive Cancer Center core grant supplement 5P30-CA-046592, NSF DMS award 1712933 and The University of Michigan precision health award U067541. We thank Alexander Rix for his work developing R package *SAMBA*. We thank Lars Fritsche and the International AMD Genomics Consortium (IAMDGC) for providing GWAS summary statistics.

## References

Allen, W. E., Altae-tran, H., Briggs, J., Jin, X., Mcgee, G., Shi, A., Raghavan, R., Kamariza, M., Nova, N., and Others, A. (2020). Population-scale longitudinal mapping of COVID-19 symptoms, behaviour and testing. Nature Human Behaviour 4, 972–982.

Beesley, L. J. and Mukherjee, B. (2020). Statistical inference for association studies using electronic health records: handling both selection bias and outcome misclassification. Biometrics.

Chadeau-Hyam, M., Bodinier, B., Elliott, J., Whitaker, M. D., Tzoulaki, I., Vermeulen, R., Kelly-irving, M., Delpierre, C., and Elliott, P. (2020). Risk factors for positive and negative COVID-19 tests: a cautious and in-depth analysis of UK biobank data. International Journal of Epidemiology pages 1–14.

Dahabreh, I. J. and Hernán, M. A. (2019). Extending inferences from a randomized trial to a target population. European Journal of Epidemiology 34, 719–722.

Elliot, M. R. (2009). Combining Data from Probability and Non-Probability Samples Using Pseudo-Weights. Survey Practice 2, 1–7.

Fritsche, L. G., Gruber, S. B., Wu, Z., Schmidt, E. M., Zawistowski, M., Moser, S. E., Blanc, V.M., Brummett, C. M., Kheterpal, S., Abecasis, G. R., and Mukherjee, B. (2018). Association of Polygenic Risk Scores for Multiple Cancers in a Phenome-wide Study: Results from The Michigan Genomics Initiative. The American Journal of Human Genetics 102, 1–14.

Fritsche, L. G., Igl, W., Cooke Bailey, J. N., Gassmann, F., Sengupta, S., Bragg-Gresham, J. J., and al, E. (2016). A large genome-wide association study of age-related macular degeneration highlights contributions of rare and common variants. Nature Genetics 48, 134–143.

Gu, T., Mack, J. A., Salvatore, M., Sankar, S. P., Valley, T. S., and Singh, K. (2020). Characteristics Associated With Racial / Ethnic Disparities in COVID-19 Outcomes in an Academic Health Care System. JAMA Network Open 3, 1–15.

Haneuse, S. and Daniels, M. (2016). A General Framework for Considering Selection Bias in EHR-Based Studies: What Data are Observed and Why? eGEMs 4, 1–17.

Huang, J., Duan, R., Hubbard, R. A., Wu, Y., Moore, J. H., Xu, H., and Chen, Y. (2018). PIE: A prior knowledge guided integrated likelihood estimation method for bias reduction in association studies using electronic health records data. Journal of the American Medical Informatics Association 25, 345–352.

Sinnott, J. A., Dai, W., Liao, K. P., Shaw, S. Y., Ananthakrishnan, A. N., Gainer, V. S., Karlson, E. W., Churchill, S., Szolovits, P., Murphy, S., Kohane, I., Plenge, R., and Cai, T. (2014). Improving the power of genetic association tests with imperfect phenotype derived from electronic medical records. Human Genetics 133, 1369–1382.

Vandenbroucke, J. P. and Pearce, N. (2019). Test-Negative Designs: Differences and Commonalities with Other Case-Control Studies and Other Patient Controls. Epidemiology 30, 838–844.

Woloshin, S., Patel, N., and Kesselheim, A. S. (2020). False negative tests for SARS-CoV-2 infection- challenges and implications. New England Journal of Medicine 383, e38.

