## Supporting Information for "Bias reduction and inference for electronic health record data under selection and phenotype misclassification: three case studies"

#### A Michigan Genomics Initiative dataset at a glance

In the main paper, we provide an overview of The Michigan Genomics Initiative (MGI) dataset and provide detailed analyses. Here, we include some additional information referred to in the main text. **Figure A.1** provides a visual schematic of the data generation mechanisms in MGI along with the difference between source and target populations. **Table A.1** provides comparisons between MGI patients, people included in the National Health and Nutrition Examination Survey (NHANES) in 2017-2018, and the US adult population. **Figure A.2** shows the rates of observed disease by age in these three groups of people, and **Table A.2** provides sources for external summary information used for these comparisons.

In this paper, we consider several EHR-derived phenotypes. International Classification of Disease (ICD) codes were used to define disease status in MGI data. These codes were aggregated into a coding system known as phenotype codes or "phecodes" following the coding systems described elsewhere (Denny et al., 2010). Cancer diagnosis was defined as receipt of any phecode corresponding to a cancer diagnosis during follow-up in the Michigan Medicine EHR. Diabetes diagnosis was defined as receipt of phecode 250 ("diabetes mellitus"), which includes diabetes types I and II. Coronary artery disease (CAD) diagnosis was defined as receipt of phecode 411.4 ("coronary atherosclerosis"). Macular degeneration diagnosis was defined as receipt of phecode 362.2 ("degeneration of macula and posterior pole of retina") among patients at least 50 years old. Since our goal in case study (b) is to study age-related macular degeneration (AMD), we will use "macular degeneration" and "AMD" interchangeably. Body mass index (BMI) was defined as the median observed BMI value prior to any cancer diagnosis or bariatric surgery. For patients without BMI measurements before such diagnoses, the earliest observed BMI was chosen.

**Figure A.1:** Schematic of MGI data generation and desired data analysis

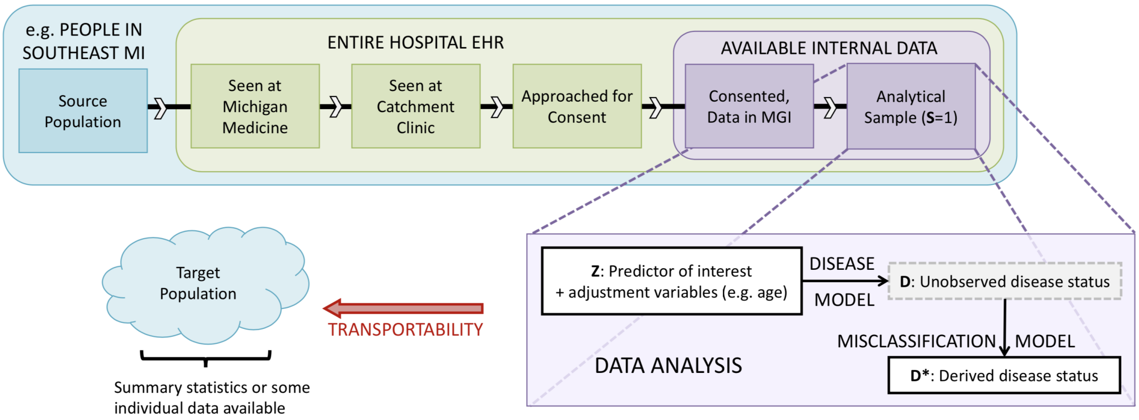

**Table A.1:** Comparison of disease and demographic characteristics between cohorts

| **Characteristic** | **MGI**[1] | **NHANES, 2017-2018 (Interview and Examination, 18+)** | **US Adult Population** |
|---|---|---|---|
| Sample size | 40101 | 5533 | >200,000,000 |
| Age, median | 59 | 51 | 35.3[3] |
| Length of follow-up (d = day, y = year) | 8.0y (1d - 40.2y) | - | - |
| Number of visits, mean (range) | 84 (1 - 1323) | - | - |
| Body Mass Index (BMI), mean | 29.9 | 29.7 | 27.6[4] |
| BMI category, n (%) | | | |
| Underweight, <18.5 | 450 (1.1) | 99 (1.8) | 11.9%[4] |
| Normal, [18.5, 25.0) | 9707 (24.2) | 1372 (24.8) | 27.3% |
| Overweight, [25.0, 30.0) | 13023 (32.5) | 1727 (31.2) | 26.3% |
| Obese, 30.0+ | 16912 (42.2) | 2236 (40.4) | 34.4% |
| Unknown | 9 (0.02) | 99 (1.8) | - |
| Female, n (%) | 1014 (52.4) | 2861 (51.7) | 50.9%[3] |
| Smoking habits, n (%) | | | |
| Never | 20003 (49.9) | 3301 (59.7) | 58.9%[4] |
| Former | 13215 (32.9) | 1260 (22.8) | 24.0% |
| Current | 6780 (16.9) | 972 (17.6) | 17.1% |
| Unknown | 103 (2.6) | 0 (0) | - |
| Lifetime disease prevalence, (%) | | | |
| Diabetes (Type I or II) | 11670 (29.1) | 838 (15.2)[2] | 13.0[5] |
| Macular degeneration | 1743 (4.3) | - | 2.1%[6] |
| Coronary artery disease | 6409 (15.6) | 243 (4.6) | 12.1%[7] |
| Cancer (any type) | 21345 (53.2) | 551 (10.5) | 39.5%[8] |

[1] age of last diagnosis in EHR.
[2] NHANES diagnosis status was missing for 4, 284, and 270 patients for diabetes, coronary artery disease, and cancer (any type), respectively. Macular degeneration diagnosis was not collected for NHANES in 2017-2018. Lifetime disease prevalence was calculated for NHANES using patients with observed disease status.
[3] Source: US Census, 2000 overall median age. Adult-only median age between 40-44.
[4] Source: National Health and Nutrition Examination Survey (NHANES) with selection weighting, 2017-2018
[5] Source: Centers for Disease Control and Prevention (CDC), National Diabetes Statistics Report, 2013-2016
[6] Source: NIH National Eye Institute Statistics, 2010 prevalence for ages 50+.
[7] Source: CDC, National Center for Health Statistics, National Health Interview Survey, 2017-2018
[8] Source: NIH National Cancer Institute; Surveillance, Epidemiology and End Results Program (SEER), 2015-2017

**Table A.2:** External data sources used for bias correction

| Data Source | Quantity | Link |
|---|---|---|
| US Census, 2000 | age distribution | https://www.census.gov |
| NIH National Cancer Institute; Surveillance, Epidemiology and End Results Program (SEER) | cancer prevalence[1] by age (2016) | https://seer.cancer.gov/data/ |
| | lifetime risk of developing cancer (all sites, 2015-2017) | https://seer.cancer.gov/data/ |
| | lifetime cancer risk by gender (all sites, 2008-2016) | https://seer.cancer.gov/csr/previous.html |
| NIH National Eye Institute Statistics, 2010 | age-related macular degeneration prevalence by age (and overall for ages 50+) | https://www.nei.nih.gov |
| Centers for Disease Control and Prevention (CDC), National Diabetes Statistics Report, 2013-2016 | diabetes prevalence by age | https://www.cdc.gov |
| CDC, National Center for Health Statistics, National Health Interview Survey, 2017-2018 | coronary artery disease prevalence by age | https://www.cdc.gov/nchs/index.htm |
| National Health and Nutrition Examination Survey (NHANES), 2017-2018 | cancer, diabetes, CAD, AMD, BMI, age, and smoking joint distribution | https://www.cdc.gov/nchs/... |

[1] invasive cancers only, limited to cancers occurring in the previous 24 years.

**Figure A.2:** Disease prevalence by age in MGI, NHANES, and the US adult population[1]

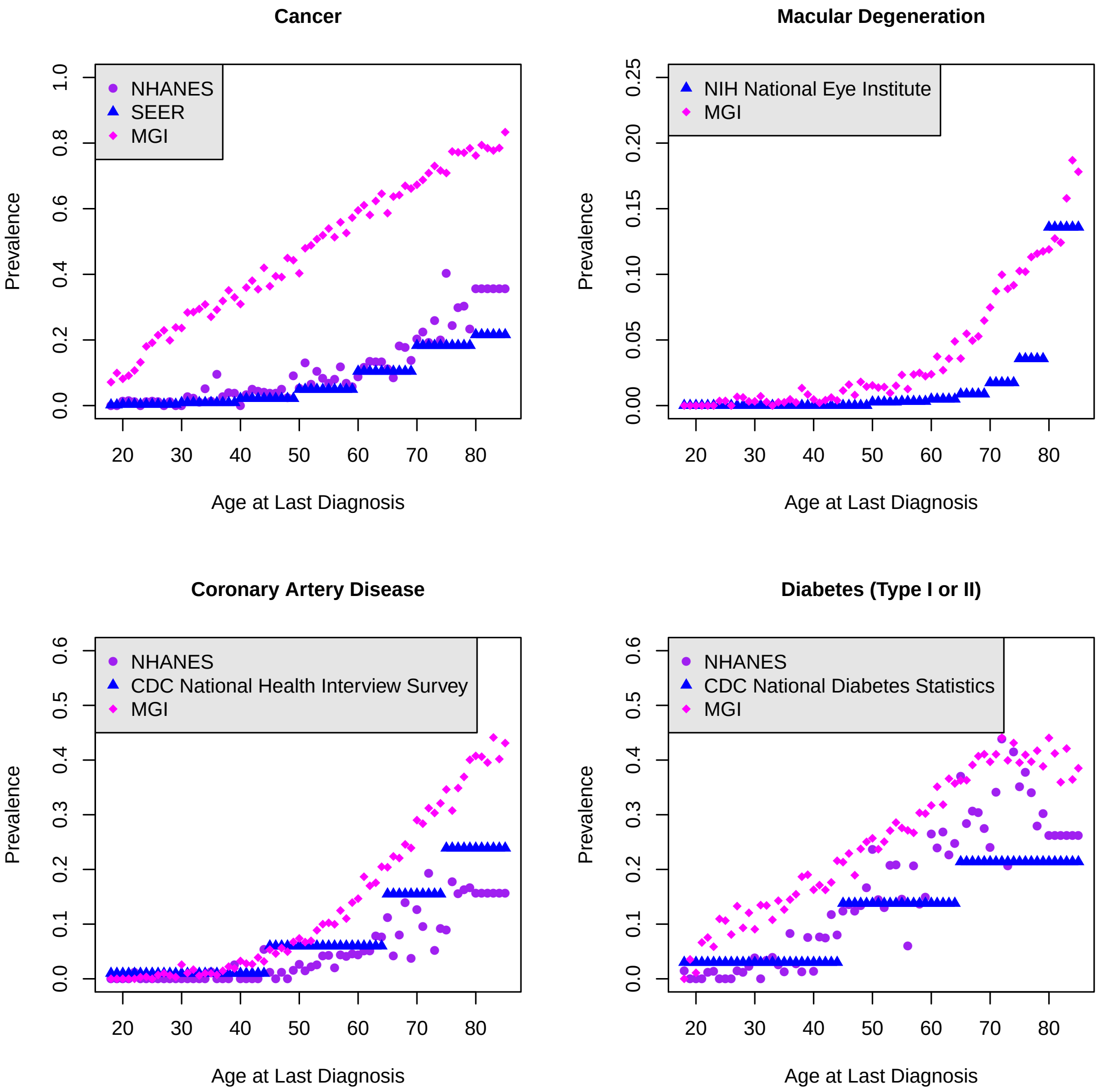

[1] Prevalence by age in NHANES was calculated using data from 2017-2018. Data on macular degeneration diagnosis are not available for NHANES for this time period. Cancer prevalence by age is for invasive cancers only.

### B SARS-CoV-2 dataset at a glance

In case study (c), we consider data for all patients tested for SARS-CoV-2 viral infection via RT-PCR in Michigan Medicine, restricted to patients who have primary care providers in Michigan Medicine. Patients were determined to be primary care patients if the patient had an encounter in any of the primary care locations within Michigan Medicine since 01-01-2018. **Table B.1** provides descriptions of these tested patients along with a simple random sample of untested controls from within Michigan Medicine. Untested controls were sampled among patients who are alive and who had an encounter with the Michigan Medicine health system since 04-23-2012.

**Table B.1:** Comparison of tested/diagnosed and control cohorts with primary care provider within Michigan Medicine[1]

| **Characteristic** | Tested/diagnosed cohort | Non-tested, non-diagnosed controls |
|---|---|---|
| Size | 9154 | 4618 |
| Number comorbidities | | |
| mean (min-max) | 2.6 (0-7) | 1.8 (0-7) |
| Unknown, n (%) | 318 (3.5) | 67 (1.5) |
| Male, n (%) | 3556 (38.8) | 2115 (45.8) |
| Age category, n (%) | | |
| <18 | 1866 (20.4) | 1191 (25.8) |
| 18-34 | 863 (9.4) | 1170 (25.3) |
| 35-49 | 1925 (21.0) | 762 (16.5) |
| 50-64 | 2232 (24.4) | 806 (17.5) |
| 65-79 | 1751 (19.1) | 551 (11.2) |
| 80+ | 517 (5.6) | 138 (3.0) |
| Smoker[2], n (%) | | |
| Current/Former | 3498 (38.2) | 1074 (23.3) |
| Never | 5380 (58.8) | 3460 (74.9) |
| Unknown | 276 (3.0) | 84 (1.8) |
| Body mass index (BMI) category[2], n (%) | | |
| Underweight, <18.5 | 122 (1.3) | 68 (1.5) |
| Healthy, [18.5, 25) | 2172 (23.7) | 1144 (24.8) |
| Overweight, [25,30) | 2449 (26.8) | 1068 (23.1) |
| Obese, 30+ | 3260 (35.6) | 1010 (21.9) |
| Unknown | 1151 (12.6) | 1328 (28.8) |
| Race/ethnicity, n (%) | | |
| Non-Hispanic White | 6769 (73.9) | 3162 (68.5) |
| Non-Hispanic African American | 1183 (12.9) | 463 (10.0) |
| Hispanic/other/multi-racial | 879 (9.6) | 764 (16.5) |
| Unknown | 323 (3.5) | 229 (5.0) |
| Neighborhood Disadvantage Index (NDI) | | |
| mean (min-max) | 0.10 (0.01-0.57) | 0.01 (0.02-0.57) |
| Unknown, n (%) | 1159 (12.7) | 513 (11.1) |
| Population Density (persons per square mile) | | |
| mean (min-max) | 2527 (10-17441) | 2535 (8-22225) |
| Unknown, n (%) | 1159 (12.7) | 513 (11.1) |
| Tested positive or diagnosed, n (%) | 710 (7.8) | - |
| Testing Quarter, n (%) | | |
| Diagnosed | 138 (1.5) | - |
| Q1 (March 10 - March 31, 2020) | 1372 (15.0) | - |
| Q2 or Q3 (April 1 - July 28, 2020) | 7644 (83.5) | - |
| Hospitalized[3], n (%) | 195 (2.1) | - |

[1] Number of comorbidities corresponds to number of disease categories (0-7) for which the patient has prior diagnoses among: respiratory diseases, circulatory diseases, any cancer, type II diabetes, kidney diseases, liver diseases, and autoimmune diseases. NDI refers to neighborhood socioeconomic disadvantage index, following Clarke et al. (2014). For additional details, refer to Gu et al. (2020).
[2] last reported value
[3] includes hospitalizations among people who later tested positive (after first test)

### C Additional figures and tables for case studies (a) and (b)

We can use the population disease rates for cancer and macular degeneration reported in **Table A.1** along with the observed EHR-derived disease status rates in MGI to obtain estimates of the marginal sampling ratio, $\tilde{r}$, as a function of potential values for the marginal sensitivity, $\tilde{c}$. **Figure C.1** shows these predicted values. We cannot use these data alone to determine the "true" value for $\tilde{r}$. Instead, we can use this plot to guide reasonable choices for $\tilde{r}$ for further analysis. For both outcomes, we consider values between 1 (no disease-related selection) and 100 (patients with disease 100x more likely to be included).

**Figure C.1:** Marginal sampling ratio as a function of marginal sensitivity

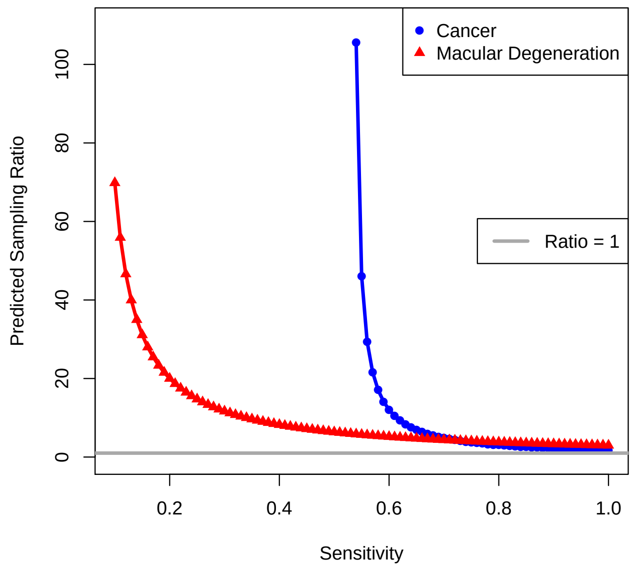

[1] These relationships were estimated using assumed disease prevalences in **Table A.1**. For the cancer outcome, some values of $\tilde{c}$ were incompatible with the data (estimated $\tilde{r} < 0$), and $\tilde{r}$ is not plotted.

We then estimate sensitivity $c_{true}(X)$ as a function of covariates $X$ using method 2b in **Figure 1**. This approach requires specification of $P(D = 1|X)$. This distribution is not known, but we do know the marginal disease prevalence, $P(D = 1)$, and the relationship between disease diagnosis and age, $P(D = 1|\text{Age})$. We estimate $c_{true}(X)$ first assuming $P(D = 1|X) = P(D = 1)$ and then assuming $P(D = 1|X) = P(D = 1|\text{Age})$. We present results assuming $P(D = 1|X) = P(D = 1|\text{Age})$ in the main paper. **Figure C.2** presents the distributions of estimated $c_{true}(X)$ across MGI participants. This figure demonstrates that the choice $P(D = 1|X)$ can have a strong impact in the estimation of $c_{true}(X)$. For the cancer outcome, where diagnosis is expected to be associated with more doctors visits and longer follow-up, neither of our specifications for $P(D = 1|X)$ is very reasonable. We showed in Beesley and Mukherjee (2020), however, that downstream estimation of $\theta$ is only weakly impacted by the specification of $P(D = 1|X)$, so we are not very concerned with this assumption violation in practice. **Figure C.3** shows the estimated $\beta$ log-odds ratios for covariates in the sensitivity model. Interestingly, the estimated odds ratios tend to be similar for the two outcomes, with a slightly stronger estimated association with longer follow-up time for the macular degeneration outcome. Higher sensitivity in both outcomes is associated with longer follow-up and more visits per follow-up time.

**Figure C.2:** Estimated patient-varying sensitivities $c_{true}(X)$ as a function of marginal sampling ratio using method 2b. $P(D=1|X)$ was assumed to equal either $P(D=1)$ or $P(D=1|\text{Age})$.

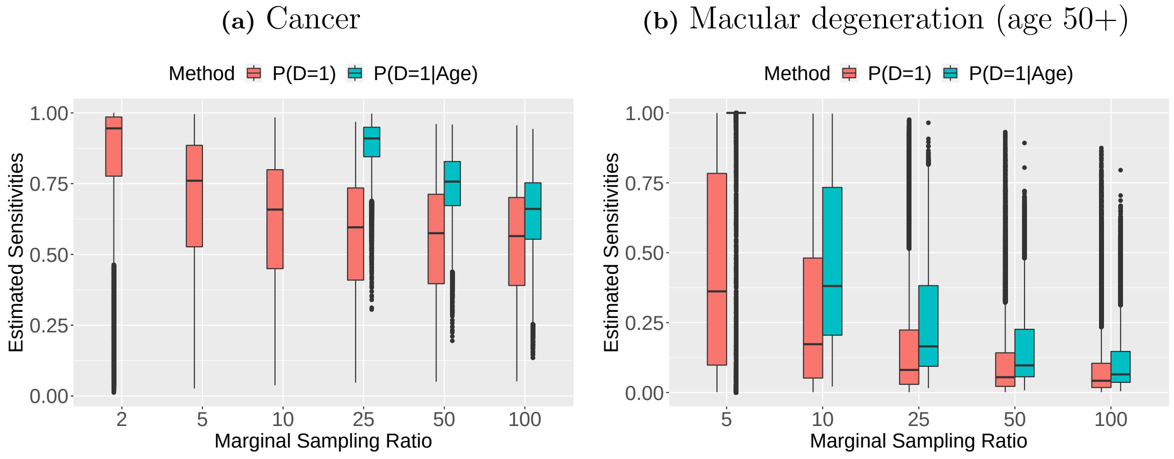

**Figure C.3:** Estimated sensitivity model parameters $\beta$ as a function of marginal sampling ratio. Sensitivity was estimated using the method 2b and setting $P(D=1|X) = P(D=1|\text{Age})$

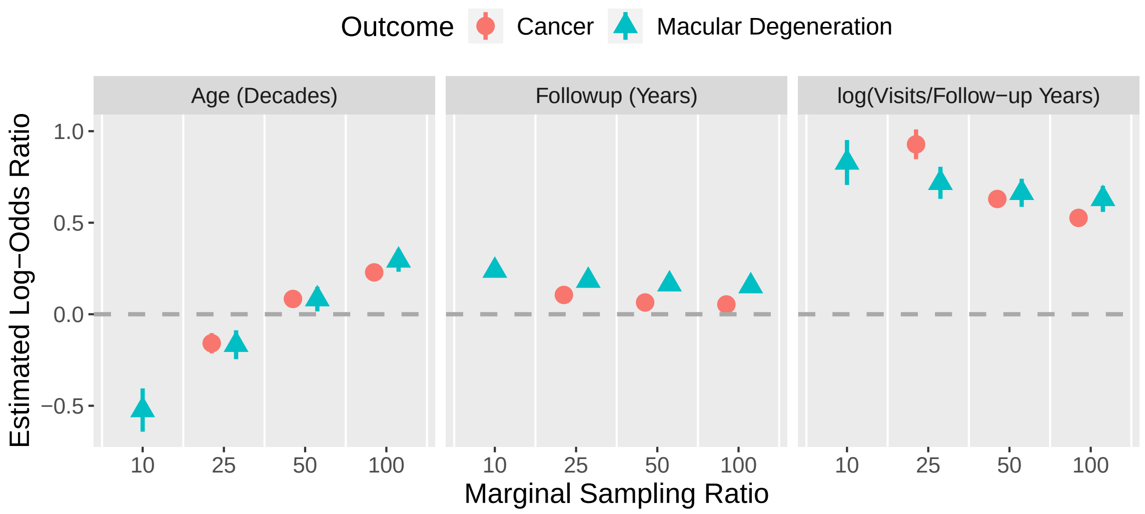

In estimating weights for selection bias adjustment using NHANES data, we fit regression models for (1) the probability of selection into NHANES among the US adult population and (2) the probability of inclusion in MGI given inclusion in MGI or NHANES. Parameter estimates for these regression model fits are provided in **Table C.1**.

**Figure C.4** shows the estimated individual-level selection weights for the 40,101 MGI patients, where each type of weight is sorted by increasing value along the x-axis. All weights are trimmed above by 10 to ensure no one patient dominates the estimation. Poststratification weights with and without the AMD outcome are extremely similar, indicating that AMD status may not appreciably differ between MGI and the US populations after accounting for differences due to age and other diseases. In contrast, poststratification weights with and without the cancer outcome differ substantially, reflecting the clear enrichment of MGI in terms of cancer outcomes relative to the US adult population. This same phenomenon occurs for weights estimated using NHANES. Additionally, the distribution of weights with and without incorporating the cancer outcome in NHANES tend to be similar to corresponding poststratification weights. This is noteworthy, since these weights are estimated using entirely different population data sources.

**Table C.1:** Beta regression of NHANES sampling probabilities (relative to US adult population) and logistic regression for including in MGI given inclusion in NHANES or MGI

| | **Beta regression for sampling probabilities**[1] Log-Odds Ratio, (95% CI) | **Logistic regression for inclusion in MGI** Log-Odds Ratio, (95% CI) |
|---|---|---|
| Age | | |
| <40 | reference | reference |
| 40-59 | 0.20 (0.15, 0.24) | 0.56 (0.48, 0.64) |
| 60+ | 0.56 (0.52, 0.61) | 0.46 (0.39, 0.54) |
| Diabetes diagnosis | | |
| No | reference | reference |
| Yes | 0.14 (0.10, 0.19) | 0.58 (0.50, 0.66) |
| CAD diagnosis | | |
| No | reference | reference |
| Yes | -0.02 (-0.09, 0.05) | 1.08 (0.94, 1.22) |
| BMI category | | |
| Underweight (<= 18.5) | 0.08 (-0.05, 0.21) | -0.29 (-0.53, -0.04) |
| Normal (18.6 – 24.9) | reference | reference |
| Overweight (25.0 – 29.9) | -0.08 (-0.12, -0.03) | -0.15 (-0.22, -0.07) |
| Obese (30+) | -0.13 (-0.18, -0.09) | -0.27 (-0.34, -0.19) |
| Smoking habits | | |
| Never | reference | reference |
| Current or Former | -0.01 (-0.05, 0.02) | - |
| Current | - | 0.06 (-0.02, 0.14) |
| Former | - | 0.29 (0.22, 0.36) |
| Race/ethnicity | | |
| Non-Hispanic White | reference | - |
| Hispanic | 0.43 (0.39, 0.47) | - |
| Non-Hispanic Black | 0.63 (0.59, 0.67) | - |
| Other (including multi) | reference | - |

[1] Implemented with a logit link function for mean of beta distribution

**Figure C.4:** Estimated selection bias adjustment weights (not correcting for misclassification of disease phenotypes)

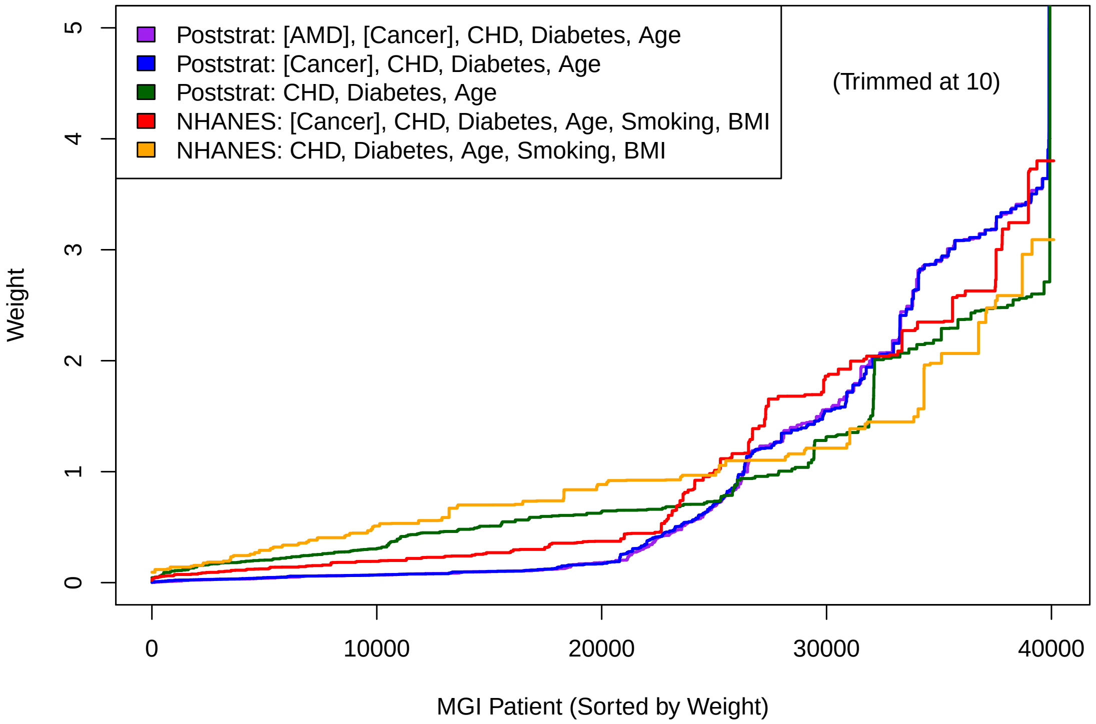

[1] "Poststrat" indicates poststratification weights estimated using population summary statistics. "NHANES" indicates IPW weights estimated using NHANES data. Estimated weights are shown along the y-axis, and each weight is sorted by increasing value across the 40101 MGI patients along the x-axis. For the AMD outcome, we show weights for all MGI patients before subsetting and rescaling to focus on the aged 50+ subset of MGI. Brackets in legend labels correspond to outcome variables for case studies (a) and (b).

**Table C.2:** Log-odds ratio point estimates and width of 95% confidence intervals for cancer-gender associations (reference = male) [Case study (a)] [1]

| | Log-odds ratio | Width of 95% confidence intervals |
|---|---|---|
| Uncorrected analysis | -0.07 | 0.079 |
| Selection weighting only | | |
| Poststratification: without cancer | 0.16 | 0.108 |
| Poststratification: with cancer (uncorrected) | -0.16 | 0.096 |
| NHANES IPW: without cancer | 0.10 | 0.095 |
| NHANES IPW: with cancer (uncorrected) | -0.11 | 0.087 |
| Approx. $D^*\|Z$ method [method 4a] | | |
| No weighting | -0.11 | 0.122 |
| Poststratification: without cancer | 0.20 | 0.138 |
| Poststratification: with cancer (uncorrected) | -0.16 | 0.098 |
| Poststratification: with cancer (corrected) | -0.15 | 0.097 |
| NHANES IPW: without cancer | 0.13 | 0.125 |
| NHANES IPW: with cancer (uncorrected) | -0.11 | 0.091 |
| NHANES IPW: with cancer (corrected) | -0.10 | 0.091 |
| Non-logistic link method [method 4c] | | |
| No weighting | -0.09 | 0.091 |
| Poststratification: without cancer | 0.16 | 0.119 |
| Poststratification: with cancer (uncorrected) | -0.17 | 0.098 |
| Poststratification: with cancer (corrected) | -0.16 | 0.098 |
| NHANES IPW: without cancer | 0.10 | 0.105 |
| NHANES IPW: with cancer (uncorrected) | -0.11 | 0.089 |
| NHANES IPW: with cancer (corrected) | -0.10 | 0.089 |

[1] For Approx. $D^*|Z$ and Non-logistic link function methods, sensitivity is estimated assuming $\tilde{r} = 25$.

**Figure C.5** compares estimated associations between macular degeneration diagnosis and 43 genetic loci in MGI and IAMDGC. The box around each point corresponds to the 95% confidence interval in either dataset. We observe smaller estimated effects in MGI compared to IAMDGC estimates. There are many possible explanations for this phenomenon, three of which seem most likely. Firstly, the 43 genetic loci were chosen as the top hits in the IAMDGC GWAS, so the resulting point estimates may be over-estimated following the "winner's" curse. Secondly, GWAS results for *advanced* AMD were obtained for IAMDGC data, and resulting genetic associations may be stronger for this outcome than for the MGI macular degeneration outcome including less advanced cases in addition to advanced ones. Thirdly, MGI results may be attenuated as a result of misclassification and/or selection bias.

**Figure C.5:** Estimated AMD log-odds for 43 SNPs using MGI and IAMDGC data. 95% confidence intervals in MGI and IAMDGC data are shown as shaded boxes.

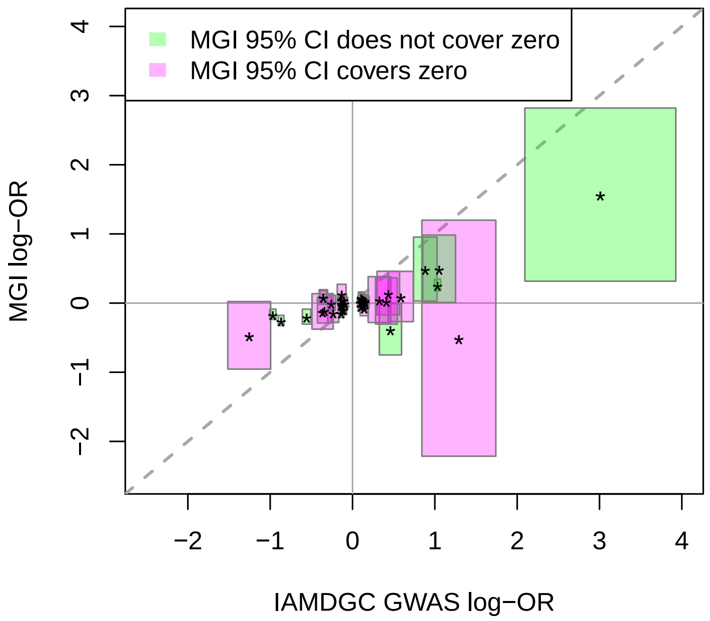

In **Table C.3**, we provide summary metrics for the performance of the bias-correction strategies in **Figure 1** to reduce potential bias due to phenotype misclassification and selection. An abridged version of this table is included and discussed in the main paper. **Figure C.6** shows the 43 estimated log-odds ratio associations in IAMDGC and MGI (uncorrected and corrected using method 4a without selection weighting). Method 4a does not uniformly map the uncorrected MGI point estimates to the IAMDGC GWAS estimates. This plot does demonstrate, however, that the point estimates and confidence intervals can sometimes differ substantially between the various data analysis methods for a given genetic locus, and these differences here are more pronounced for extreme values of the IAMDGC GWAS $\theta$ (far left and far right values).

**Figure C.6:** Bias-adjusted AMD log-odds for estimates for 43 SNPs

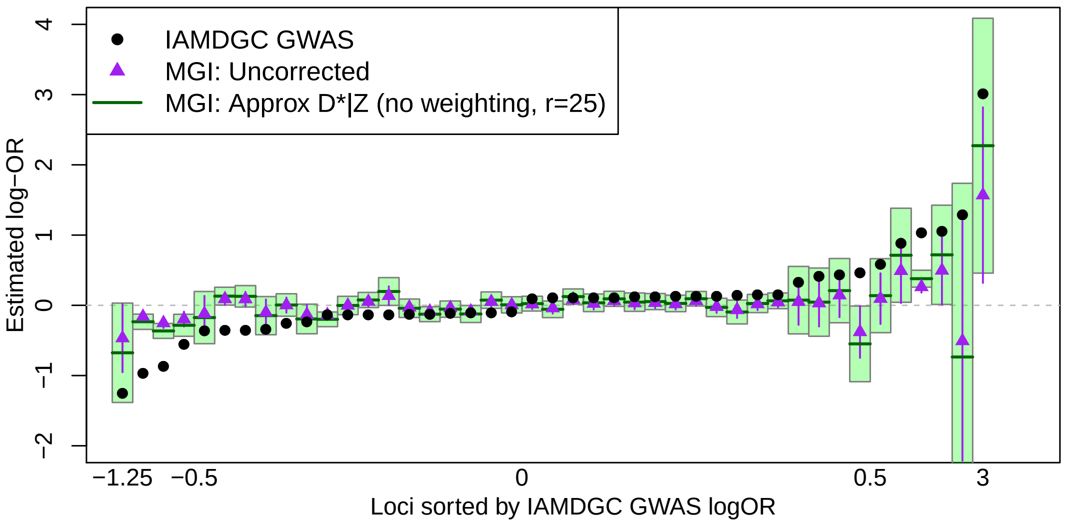

**Table C.3:** Bias-adjusted AMD log-odds ratios across 43 genetic loci [Case study (b)] [1]

| | **Avg. Absolute Deviation** | **Lin's Concordance Correlation** | **MAPE** | **Avg. Relative Standard Error** |
|---|---|---|---|---|
| IAMDGC GWAS | 0 | 1 | 0 | 1 |
| Uncorrected Analysis | 0.32 | 0.55 | **0.84** | 2.3 |
| No misclassification adjustment | | | | |
| Weighting without AMD | **0.27** | **0.68** | **0.87** | 4.4 |
| Weighting with AMD (Uncorrected) | 0.33 | **0.68** | 1.02 | 6.5 |
| Approx. $D^*\|Z$ method [method 4a] | | | | |
| No weighting | **0.29** | **0.67** | **0.81** | 3.3 |
| Weighting without AMD | **0.30** | **0.70** | 0.93 | 5.8 |
| Weighting with AMD (Uncorrected) | 0.34 | **0.68** | 1.07 | 7.2 |
| Weighting with AMD (Marg. Corrected) | 0.33 | **0.68** | 1.03 | 6.9 |
| Non-logistic link method [method 4c] | | | | |
| No weighting | **0.31** | 0.61 | **0.84** | 3.2 |
| Weighting without AMD | **0.31** | 0.33 | 1.02 | 5.7 |
| Weighting with AMD (Uncorrected) | 0.38 | **0.68** | 1.21 | 7.5 |
| Weighting with AMD (Corrected) | 0.33 | **0.69** | 1.08 | 6.9 |

[1] For Approx. $D^*|Z$ and Non-logistic link function methods, sensitivity is estimated assuming $\tilde{r} = 25$. Bolded values indicate the best performing methods.
Average absolute deviation = average absolute difference between MGI and IAMDGC point estimates (lower is better)
**Definitions:** Average absolute deviation = average absolute difference between MGI and IAMDGC point estimates (lower is better); Lin's concordance correlation = estimated concordance between MGI and IAMDGC point estimates (higher is better); MAPE (mean absolute percentage error) = average absolute difference between 1 and the ratio of MGI and IAMDGC point estimates (lower is better); Avg. relative standard error = ratio of standard errors for MGI and IAMDGC point estimates.

### D Additional figures and tables for case study (c)

We want to leverage data from 4618 non-tested controls to estimate selection/testing weights to use for estimating parameters in the disease/infection model. However, missing data present a challenge to this estimation. While missingness tends to be small for any one variable, the number of people with missingness in at least one variable is larger (25% for tested patients and 40% for untested controls). We handle missing covariate information using multiple imputation by chained equations (MICE) under assumptions that missingness is independent of unobserved factors (missing at random). We perform this imputation separately for the tested and untested cohorts. We specify the following regression model structures for imputing each covariate with missingness:

- smoking and ethnicity (Hispanic vs not): logistic regression
- neighborhood disadvantage index and log10(pop. density+1): predictive mean matching
- race (White, African American, other) and BMI category: multinomial regression
- comorbidity score: ordered proportional odds regression

We obtained 10 imputed datasets. All subsequent analyses were performed separately for each of the 10 imputed datasets, and final outcome parameter estimates were combined using Rubin's multiple imputation combining rules.

For each imputed dataset, we model whether or not patients were tested using the combined tested and untested control datasets. Due to the changes in testing requirements and availability over calendar time, we consider testing within quarters of calendar time, defined as follows: Q1 = March 10th - March 31st, Q2 = April 1st - June 30th, and Q3 = July 1st - July 28th, 2020. Using the merged tested ($S = 1$) and untested ($S_{ext} = 1$) datasets, we model selection in $Q1$ and $Q2/Q3$ separately, where the latter model conditions on not being tested in $Q1$. We define the following weights:

$$\omega_0 \propto \frac{1 - P(S = 1|W, S_{ext} = 1 \text{ or } S = 1)}{P(S = 1|W, S_{ext} = 1 \text{ or } S = 1)}$$

where

$$\begin{aligned} &P(S = 1|W, S_{ext} = 1 \text{ or } S = 1) \\ &\quad = 1 - P(Q1 = 0|W, S_{ext} = 1 \text{ or } S = 1)P(\text{Q2=1 or Q3=1}|Q1 = 0, W, S_{ext} = 1 \text{ or } S = 1) \end{aligned}$$

and where $Q1$, $Q2$, and $Q3$ are indicators for whether the first RT-PCR viral test occurred in the corresponding quarter. Using available data on age, race/ethnicity, gender, comorbidities, BMI, smoking habits, neighborhood disadvantage index, and population density as $W$, we estimate these selection probabilities and corresponding $\omega_0$ in each imputed dataset as summarized in **Table D.1** and **Figure D.1**.

We then fit unweighted and weighted regression models for test positivity among the tested cohort. Results are shown in **Figure D.2**. To facilitate evaluation of the potential for selection bias, we also provide results from logistic regression modeling of either positive or negative test results, using untested patients as controls. We also compare results from modeling hospitalization among people testing positive with or without selection weighting in **Figure D.3**.

**Table D.1:** Results from logistic regression modeling of viral testing or external diagnosis by quarter

| Characteristic | Testing in Q1<br>logOR (95% CI) | Testing in Q2 or Q3 or external diagnosis given no testing in Q1<br>logOR (95% CI) |
|---|---|---|
| Age category | | |
| <18 | -0.92 (-1.15, -0.68) | -0.45 (-0.58, -0.33) |
| 18-34 | reference | reference |
| 35-49 | 0.09 (-0.07, 0.25) | 0.20 (0.08, 0.32) |
| 50-64 | -0.30 (-0.47, -0.12) | 0.20 (0.07, 0.32) |
| 65-79 | -0.83 (-1.05, -0.61) | 0.26 (0.12, 0.40) |
| 80+ | -0.75 (-1.08, -0.42) | 0.40 (0.18, 0.62) |
| Race/ethnicity | | |
| Non-Hispanic White | reference | reference |
| Non-Hispanic African American | 0.40 (0.22, 0.58) | -0.02 (-0.16, 0.12) |
| Hispanic/other/multi-racial | -0.06 (-0.25, 0.13) | -0.43 (-0.55, -0.31) |
| Male | -0.29 (-0.41, -0.17) | -0.24 (-0.32, -0.16) |
| Number comorbidities | | |
| 0 | reference | reference |
| 1 | -0.01 (-0.25, 0.24) | -0.27 (-0.41, -0.13) |
| 2 | 0.31 (0.08, 0.55) | 0.08 (-0.06, 0.23) |
| 3+ | 0.47 (0.23, 0.71) | 0.64 (0.49, 0.79) |
| BMI category | | |
| Underweight/Healthy, [0-25) | reference | reference |
| Overweight [25,30) | 0.03 (-0.12, 0.19) | 0.14 (0.04, 0.23) |
| Obese 30+ | 0.17 (0.02, 0.32) | 0.35 (0.24, 0.45) |
| NDI x 10 | -0.07 (-0.15, 0.01) | 0.11 (0.05, 0.17) |
| Never smoker | 0.14 (0.01, 0.27) | -0.34 (-0.43, -0.25) |
| log10(population density+1) | 0.08 (-0.04, 0.19) | 0.00 (-0.08, 0.08) |

[1] NDI = Neighborhood disadvantage index. Results have been aggregated across 10 imputed datasets using Rubin's combining rules.

**Figure D.1:** Estimated merged-quarter selection weights for tested patients across 10 imputed datasets[1]

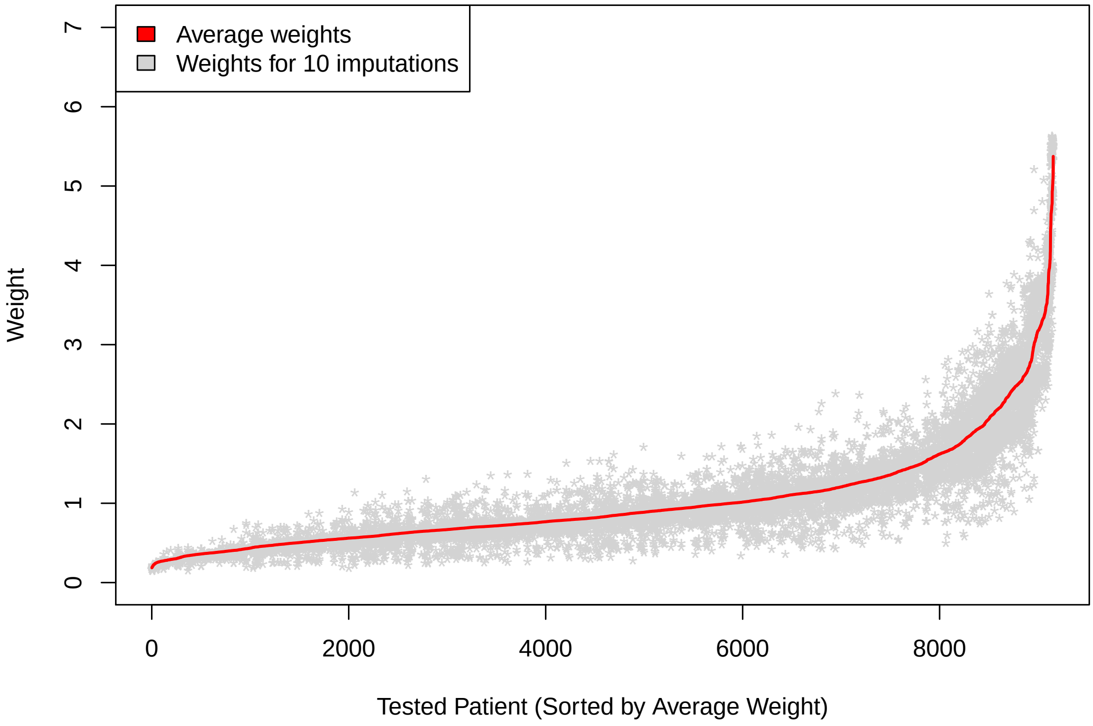

1

**Figure D.2:** Associations between patient characteristics and coronavirus infection rates with and without accounting for testing through weighting[1]

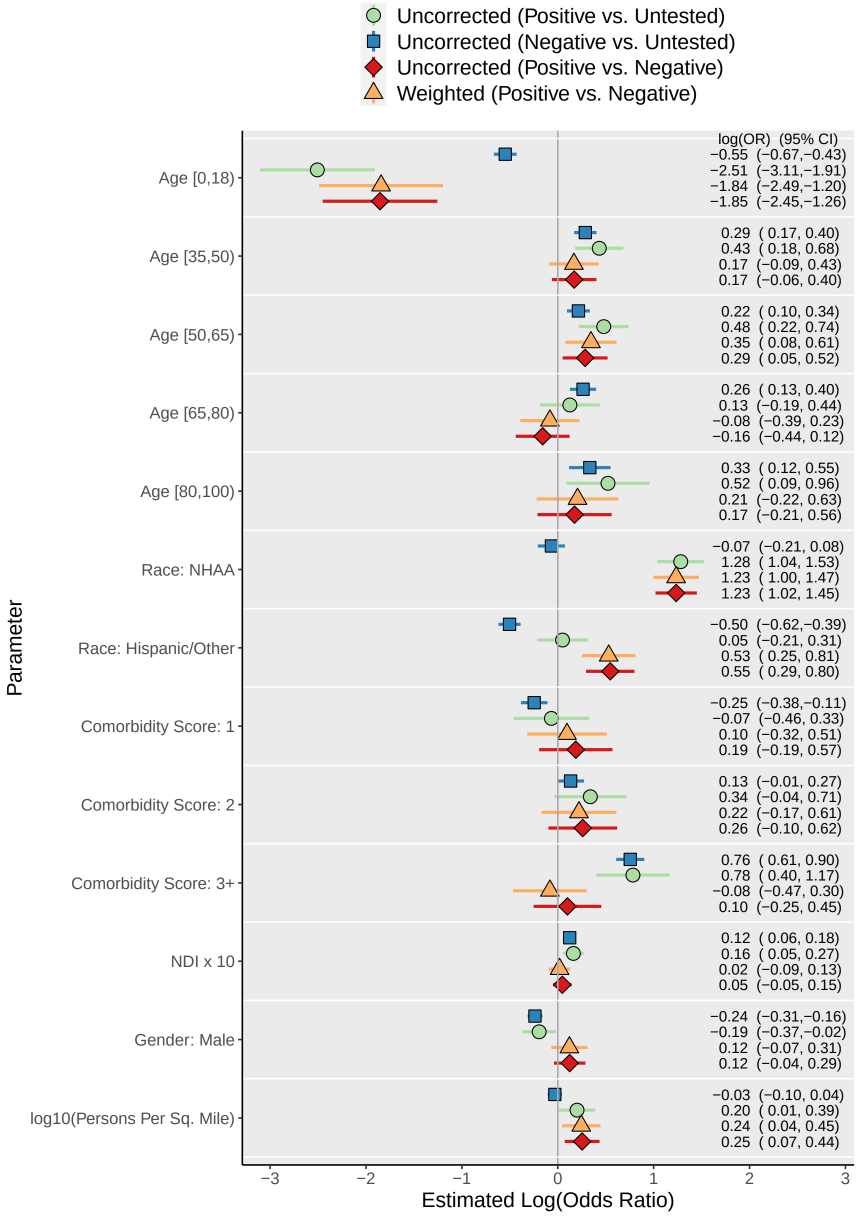

[1] NDI = Neighborhood disadvantage index. Parentheses in the legend labels correspond to the binary outcome comparison. For example, negative vs. untested corresponds to a model for testing (yes/no) excluding test-positive patients. The log-odds ratio estimate and 95% confidence interval is printed to the right of each plotted value.

**Figure D.3:** Associations between patient characteristics and hospitalization rates among people testing positive with and without accounting for testing through weighting[1]

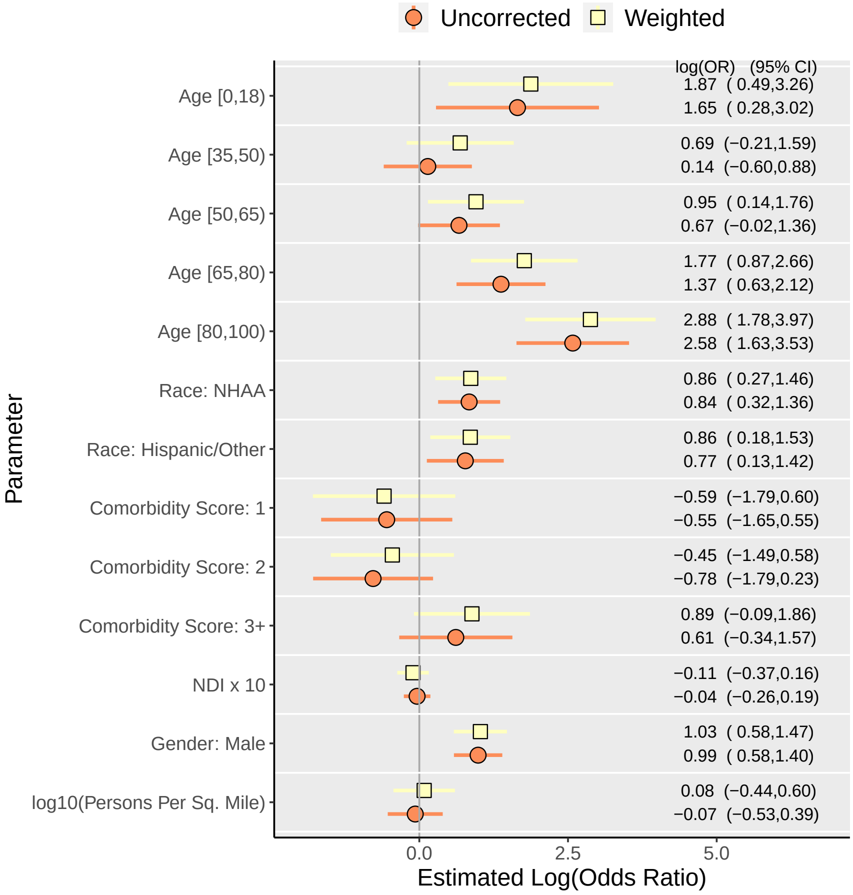

[1] NDI = Neighborhood disadvantage index. The log-odds ratio estimate and 95% confidence interval is printed to the right of each plotted value.

#### References

Lauren J Beesley and Bhramar Mukherjee. Statistical inference for association studies using electronic health records: handling both selection bias and outcome misclassification. *Biometrics*, 2020.

Philippa Clarke, Jeffrey Morenoff, Michelle Debbink, Ezra Golberstein, Michael R Elliott, and Paula M Lantz. Cumulative Exposure to Neighborhood Context : Consequences for Health Transitions Over the Adult Life Course. *Research on Aging*, 36(1):115–142, 2014. doi: 10.1177/0164027512470702.

Joshua C. Denny, Marylyn D. Ritchie, Melissa A. Basford, Jill M. Pulley, Lisa Bastarache, Kristin Brown-Gentry, Deede Wang, Dan R. Masys, Dan M. Roden, and Dana C. Crawford. PheWAS: demonstrating the feasibility of a phenome-wide scan to discover gene-disease associations. *Bioinformatics*, 26(9):1205–1210, 2010.

Tian Gu, Jasmine A Mack, Maxwell Salvatore, Swaraaj Prabhu Sankar, Thomas S Valley, and Karandeep Singh. Characteristics Associated With Racial / Ethnic Disparities in COVID-19 Outcomes in an Academic Health Care System. *JAMA Network Open*, 3(10):1–15, 2020. doi: 10.1001/jamanetworkopen.2020.25197.
